# Autoantibodies in COVID-19 correlate with anti-viral humoral responses and distinct immune signatures

**DOI:** 10.1101/2022.01.08.22268901

**Authors:** Patrick Taeschler, Carlo Cervia, Yves Zurbuchen, Sara Hasler, Christian Pou, Ziyang Tan, Sarah Adamo, Miro E. Raeber, Esther Bächli, Alain Rudiger, Melina Stüssi-Helbling, Lars C. Huber, Petter Brodin, Jakob Nilsson, Elsbeth Probst-Müller, Onur Boyman

**Author notes:** Corresponding author: Onur Boyman, MD, Department of Immunology, University Hospital Zurich, Schmelzbergstrasse 26, 8091 Zurich, Switzerland. Contributed equally (Patrick Taeschler, Carlo Cervia).

## Abstract

**Background:** Several autoimmune features occur during coronavirus disease 2019 (COVID-19), with possible implications for disease course, immunity, and autoimmune pathology. In this study, we longitudinally screened for clinically relevant systemic autoantibodies to assess their prevalence, temporal trajectory, and association with immunity, comorbidities, and severity of COVID-19.

**Methods:** We performed highly sensitive indirect immunofluorescence assays to detect anti-nuclear antibodies (ANA) and anti-neutrophil cytoplasmic antibodies (ANCA), along with serum proteomics and virome-wide serological profiling in a multicentric cohort of 175 COVID-19 patients followed-up to one year after infection, eleven vaccinated individuals, and 41 unexposed controls.

**Results:** Compared to healthy controls, similar prevalence and patterns of ANA were present in patients during acute COVID-19 and recovery. However, paired analysis revealed a subgroup of patients with transient presence of certain ANA patterns during acute COVID-19. Furthermore, patients with severe COVID-19 exhibited a high prevalence of ANCA during acute disease. These autoantibodies were quantitatively associated with higher SARS-CoV-2-specific antibody titers in COVID-19 patients and in vaccinated individuals, thus linking autoantibody production to increased antigen-specific humoral responses. Notably, the qualitative breadth of antibodies cross-reactive with other coronaviruses was comparable in ANA-positive and ANA-negative individuals during acute COVID-19. In autoantibody-positive patients, multiparametric characterization demonstrated an inflammatory signature during acute COVID-19 and alterations of the B cell compartment after recovery.

**Conclusion:** Highly sensitive indirect immunofluorescence assays revealed transient autoantibody production during acute SARS-CoV-2 infection, while the presence of autoantibodies in COVID-19 patients correlated with increased anti-viral humoral immune responses and inflammatory immune signatures.

## Introduction

Acute coronavirus disease 19 (COVID-19) causes a large clinical spectrum, ranging from a mild condition in the majority of cases to fatal disease in 1-2% of subjects.^1-3^ Several features of acute COVID-19 resemble clinical manifestations of systemic inflammatory and autoimmune diseases, such as fatigue, myalgia, hyperinflammation, thrombosis, and skin rashes.^3,4^ Furthermore, COVID-19 may trigger the onset of autoimmune pathology, as reported for Guillain-Barré syndrome, anti-phospholipid syndrome, vasculitis, and multisystem inflammatory syndrome in children.^5-9^ *Vice versa*, autoimmune phenomena have been connected to the pathogenesis of severe COVID-19. Pre-existing autoantibodies targeting the type I interferon pathway have been found in about 10% of COVID-19 cases with critical disease.^10-12^

Other acute or chronic viral infections have been associated with autoimmune responses, which have been proposed to arise by molecular mimicry, epitope spreading or bystander activation.^13^ Various autoantibodies have been described in association with COVID-19, including anti-nuclear antibodies (ANA),^14-20^ anti-neutrophil cytoplasmic antibodies (ANCA),^15,16,21^ anti- phospholipid antibodies,^5,8,14,17,19,22^ and antibodies targeting different extracellular antigens.^11,16^ While the presence of different autoantibodies has been associated with severe COVID-19 and worse outcome,^11,15,17-19^ it remains unclear to what extent autoantibodies are triggered by acute infection, even though transient autoreactivity and new development of autoantibodies have been suggested in a subgroup of COVID-19 patients.^16,20^ Furthermore, several aspects of autoantibodies in COVID-19, including their interplay with virus-specific humoral responses and their durability after acute infection, need further elucidation. In this study, we comprehensively characterized autoantibodies by using highly sensitive indirect immunofluorescence (IIF) assays in a multicentric prospective cohort of 227 individuals.

## Results

### Presence of systemic autoantibodies during acute COVID-19 and recovery

We performed a comprehensive immunological characterization of 175 individuals with confirmed COVID-19 up to one year after infection (**Fig. 1A** and **Table 1, Table S1**), including autoantibody screening by IIF, serum proteomics and serological profiling. 41 individuals with negative history and serology for severe acute respiratory syndrome coronavirus 2 (SARS-CoV-2) infection were included as controls (**Fig. 1A** and **Table 1**). Furthermore, eleven unexposed individuals were sampled before and after vaccination with BNT162b2 (**Table S2**). Using a highly sensitive IIF screening assay, we detected titers of 1:320 and above in 17 of 41 (41.4%) healthy individuals thus testing positive for ANA (**Fig. 1B**). This prevalence of ANA positivity was similar to that in COVID-19 patients during acute disease (48.0%, odds ratio (OR) = 1.30, p = 0.49) as well as six months (47.4%, OR = 1.27, p = 0.59) and one year after recovery (42.3%, OR = 1.04, p = 1) (**Fig. 1C**–**E**). Most of the observed ANA titers were just above the positivity threshold of 1:320. Interestingly, we observed a trend of higher ANA prevalence in individuals with severe COVID-19 compared to mild disease during acute infection (OR 1.85, p = 0.061), which was significantly higher at six months after recovery (OR = 3.81, p = 0.0015) (**Fig. 1C** and **D**).

**Figure 1.**
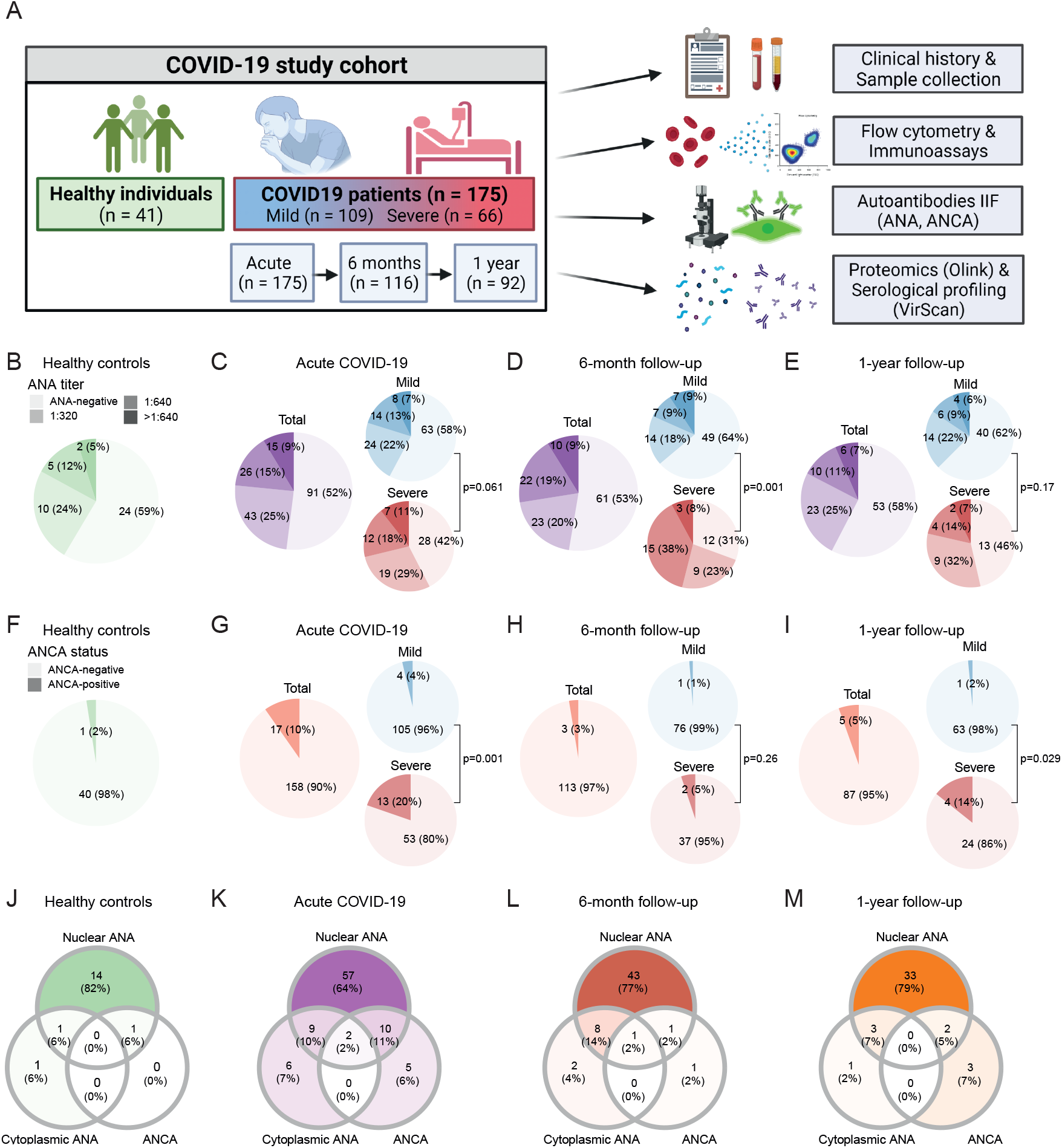
Prevalence of autoantibodies in healthy controls and COVID-19 patients during acute disease and follow-up. (**A**) Study overview. (**B**–**I**) Prevalence of ANA titers (B-E) and ANCA (F-I) in healthy controls (n = 41) and COVID-19 patients during acute disease (n = 175), six months (n = 116) and one year (n = 92) after symptom onset. (**J**–**M**) Venn diagrams depicting co-occurrence of nuclear ANA, cytoplasmic ANA and ANCA in healthy individuals (J; n = 17), acute COVID-19 patients (K; n = 89) and COVID-19 patients six months (L; n = 56) or one year (M; n = 42) after SARS-CoV-2 infection that presented with at least one type of autoantibody. P-values indicate comparison of ANA (B-E) and ANCA (F-I) prevalence between mild and severe COVID-19 patients using Fisher’ s exact test.

**Table 1.**
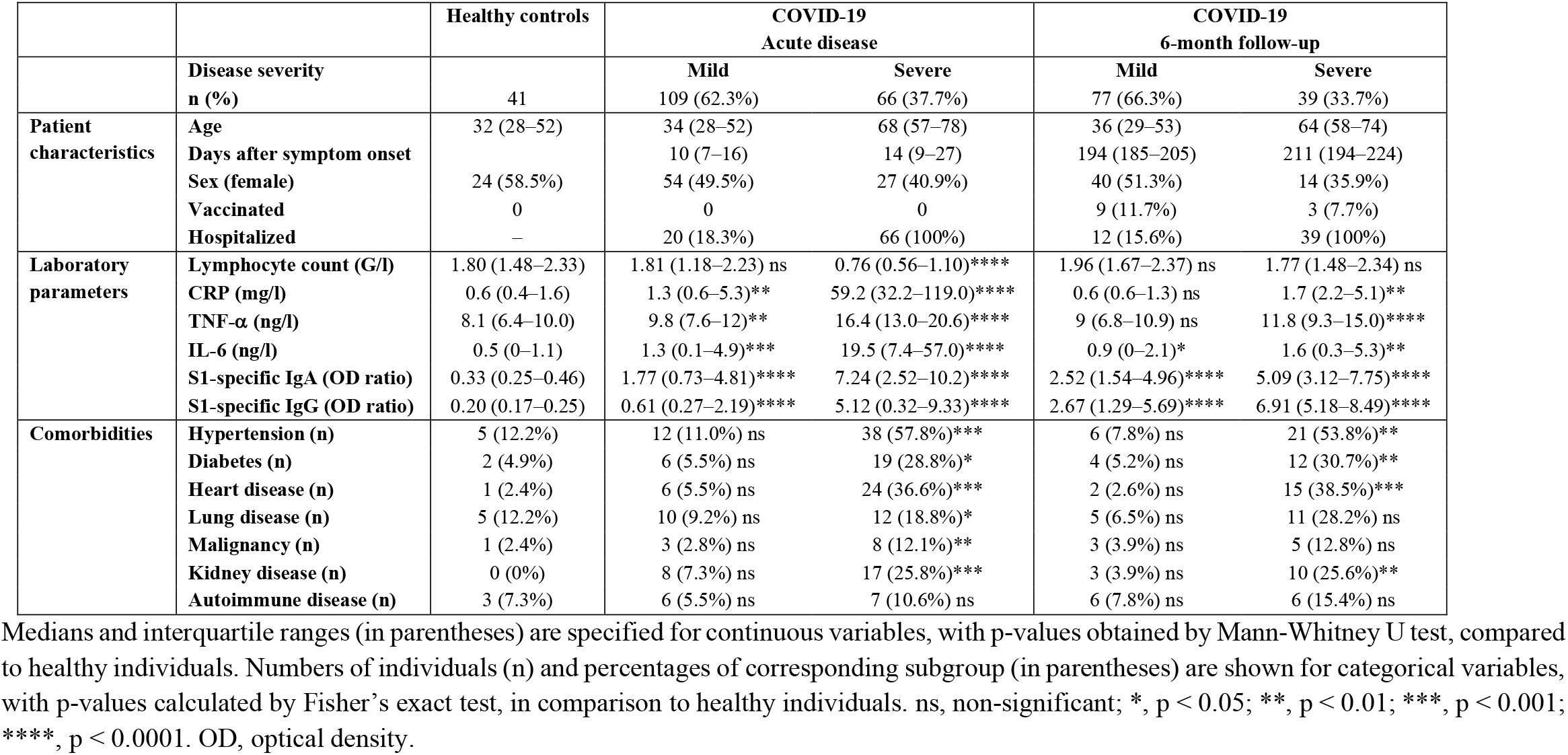
COVID-19 study cohort characteristics.

Similarly, we used an IIF assay to detect ANCA. ANCA prevalence was similar in mild COVID-19 patients during acute disease (3.6%) compared to healthy individuals (2.4%) (**Fig. 1F** and **G**). Conversely, we observed a significantly higher ANCA prevalence in severe acute COVID-19 patients (19.7%), both compared to healthy subjects (p = 0.0082) and mild COVID- 19 cases (p = 0.0096) (**Fig. 1F** and **G**), which returned to ranges seen in healthy individuals after six months (5.1%, p = 0.61) and one year (14.3%, p = 0.15) (**Fig. 1H** and **I**). In several patients, nuclear ANA, cytoplasmic ANA, or ANCA were detected concurrently, particularly during acute COVID-19 (**Fig. 1J**–**M**). Moreover, ANCA showed a tendency to be more frequent in ANA-positive (14.3%) compared to ANA-negative (5.5%) individuals during acute COVID-19 (p = 0.06) (**Fig. 1K**).

### Characteristics of ANA and ANCA patterns in acute COVID-19

To gain a qualitative appreciation, we classified ANA patterns according to the international consensus on ANA patterns anti-cell (AC) nomenclature.^23^ ANA patterns were very similar in healthy controls and COVID-19 patients at all three sampling timepoints, and in some participants different patterns were detected concurrently (**Fig. 2A**–**D, Fig. S1A**–**D**). The most common nuclear patterns were fine-granular nuclear (AC-4 or AC-4 like) and nucleolar (AC- 8, AC-9, and AC-10), whereas the most common cytoplasmic patterns were speckled (AC-19 and AC-20) (**Fig. 2E**–**G**).

**Figure 2.**
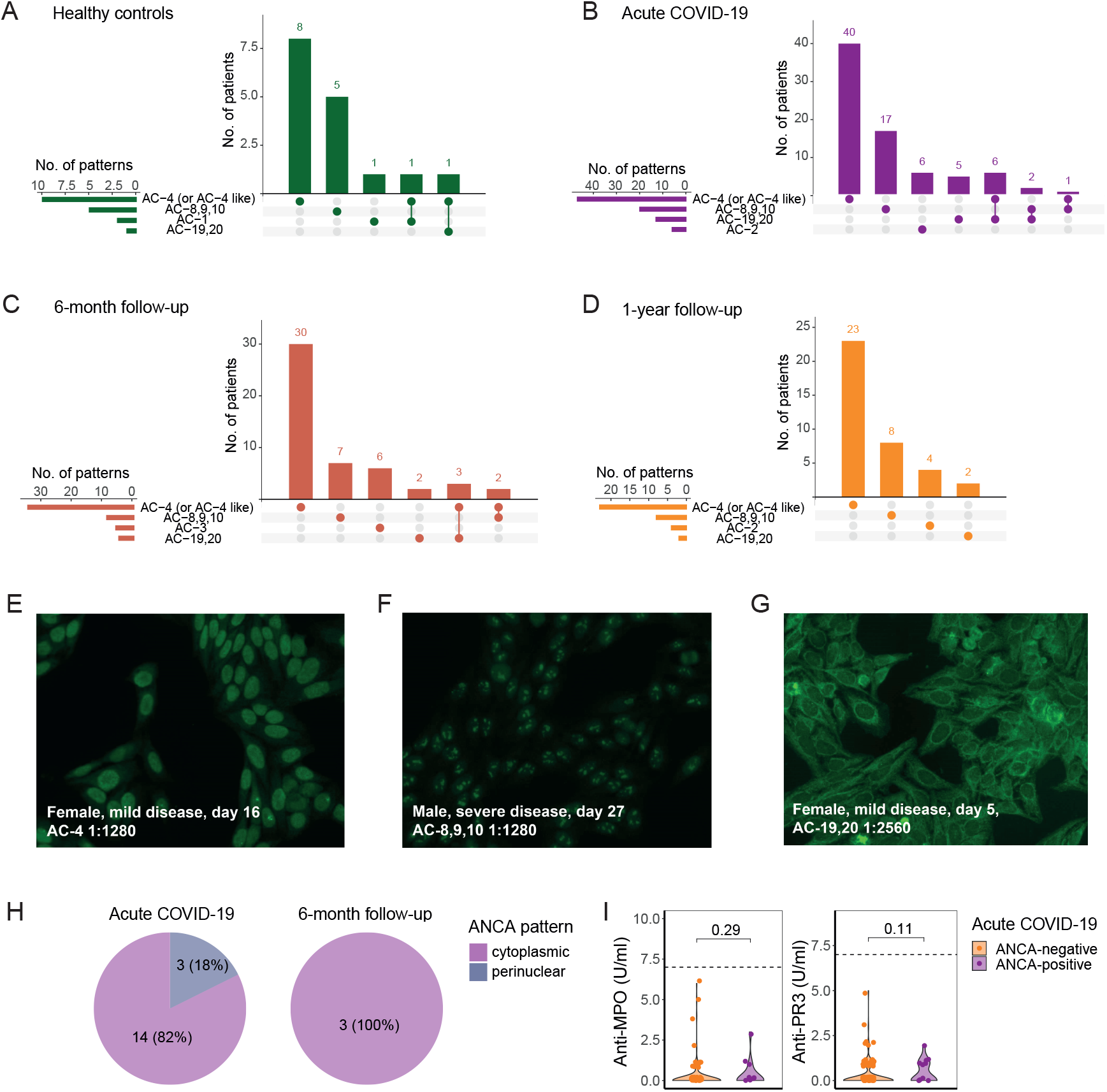
IIF pattern of autoantibodies in acute and recovered COVID-19. **(A**–**D)** Intersection plots showing counts of the four most prevalent ANA patterns (horizontal bars) and counts of pattern combinations (vertical bars) as indicated by the dot matrix, for healthy controls (A), and COVID-19 patients during acute disease (B), six months (C) and one year after symptom onset (D). (**E**–**G**) Example IIF pictures showing the most common nuclear, including fine-granular (E) and nucleolar (F), and cytoplasmic, including speckled (G), ANA patterns observed in the study cohort. All images were recorded at a dilution of 1:320. y/o, years old. (**H**) IIF ANCA patterns observed in ANCA-positive COVID-19 patients during acute disease (n = 17) and six months after recovery (n = 3). (**I**) Anti-MPO and anti-PR3 antibodies during acute COVID-19 (n = 175) in ANCA-positive and ANCA-negative individuals. Dashed lines indicate diagnostic cut-off values.

ANCA patterns observed during acute COVID-19 and follow-up were mostly cytoplasmic (**Fig. 2H**). However, cytoplasmic patterns were atypical and, accordingly, none of the ANCA-positive patients showed positivity for either anti-myeloperoxidase (MPO) or anti-proteinase 3 (PR3) antibodies (**Fig. 2I**), suggesting other antigen specificities than commonly found in ANCA-associated vasculitis ^24,25^.

### Temporal trajectory of autoantibodies in individual COVID-19 patients

To appreciate changes in ANA and ANCA on an individual level, we performed paired analysis of all followed-up COVID-19 patients (n = 129). For mild and severe COVID-19 patients combined, we observed similar proportions of patients with isolated ANA positivity during acute disease (14.7%) and follow-up (12.4%). However, a trend toward a higher proportion of new ANA development at follow-up visit was evident in patients with severe COVID-19 (20.5%) compared to patients with mild COVID-19 (8.2%) (OR = 2.84, p = 0.054) (**Fig. 3A** and **B**). To also account for subtle changes of IIF patterns, we conducted a blinded, paired analysis of IIF images to identify patterns that were transiently present either during acute disease or follow-up. Strikingly, we found transient patterns in 11 of 62 (17.7%) ANA-positive COVID-19 patients during acute disease, with speckled cytoplasmic (AC-19 and 20), nucleolar (AC-8, 9, 10) and mitotic being the most frequent patterns (**Fig. 3C** and **D**). In stark contrast, only three of 59 (5.1%) ANA-positive individuals presented with a pattern during follow-up that was not present during acute disease, thus demonstrating that transient ANA patterns were significantly more prevalent during acute COVID-19 (p = 0.045) (**Fig. 3C**).

**Figure 3.**
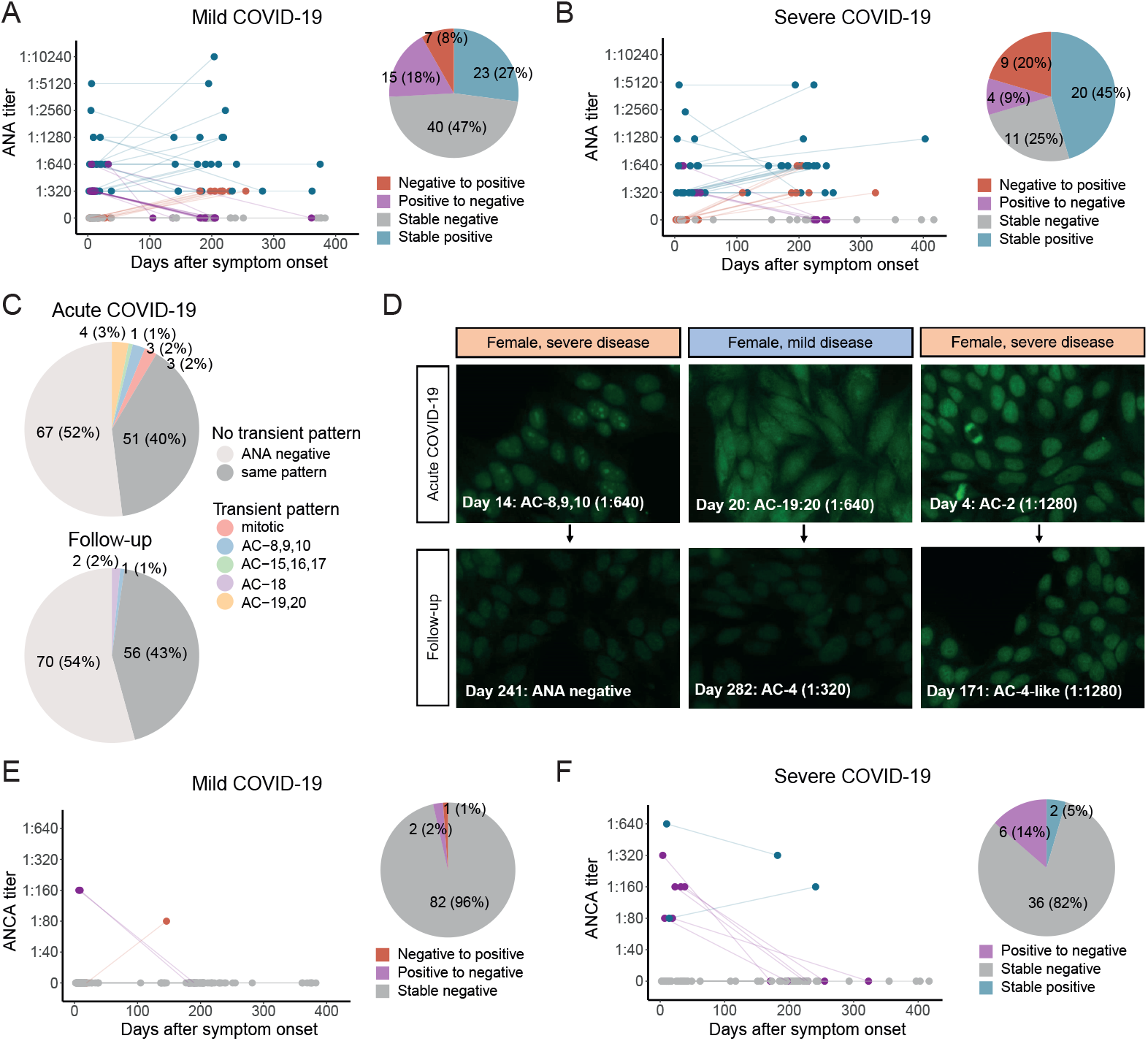
Paired longitudinal comparison indicates transient induction of autoantibodies in acute COVID-19. (**A**–**B**) Temporal trajectory of ANA titers in mild (A, n = 85) and severe (B, n = 44) COVID-19 patients, showing the first available follow-up sample, i.e. at six months (n = 116) or one year (n = 13) after symptom onset. Colors indicate development of ANA status from acute disease to follow-up. (**C**) Results from blinded, paired IIF picture analysis (n = 129). Patterns that were uniquely observed at one timepoint are colored. (**D**) Exemplary IIF pictures of three patients exhibiting transient ANA patterns during acute COVID-19, with a transient nucleolar (left), cytoplasmic (middle), or mitotic (right) pattern. All pictures were recorded at a dilution of 1:320. y/o, years old. (**E**–**F**) Temporal trajectory of ANCA titers in mild (E, n = 85) and severe (F, n = 44) COVID-19 patients. Colors indicate development of ANCA status from acute disease to follow-up.

For ANCA, we observed that of ten patients that tested positive during acute COVID-19, eight were negative during follow-up, whereas only two remained positive. Furthermore, only one patient newly exhibited positive ANCA at follow-up (**Fig. 3E** and **F**). Collectively, we found that a subgroup of individuals shows ANA and atypical ANCA production during the acute phase of COVID-19, which usually subsides during follow-up.

### Virus-specific responses in autoantibody-positive and SARS-CoV-2-vaccinated subjects

To elucidate the influence of autoantibody production during acute infection, we investigated the correlation of autoantibodies with specific humoral immune responses to SARS-CoV-2. We longitudinally assessed SARS-CoV-2 spike 1 (S1)-specific immunoglobulin A (IgA) and IgG titers and found that presence of ANA was associated with higher concentrations of S1- specific antibodies in COVID-19 patients during acute disease, which extended to six months after recovery (**Fig. 4A**). Conversely, one year after recovery, we did not observe any differences (**Fig. S2**). The presence of ANA correlated significantly with S1-specific IgG levels even after accounting for age, disease severity, and sampling timepoint in a multiple linear regression model, which was not the case of S1-specific IgA (**Fig. 4B, Table S3**). Similarly, we found higher S1-specific IgA and a trend toward higher IgG titers in patients that tested positive for ANCA during acute disease (**Fig. 4C**).

**Figure 4.**
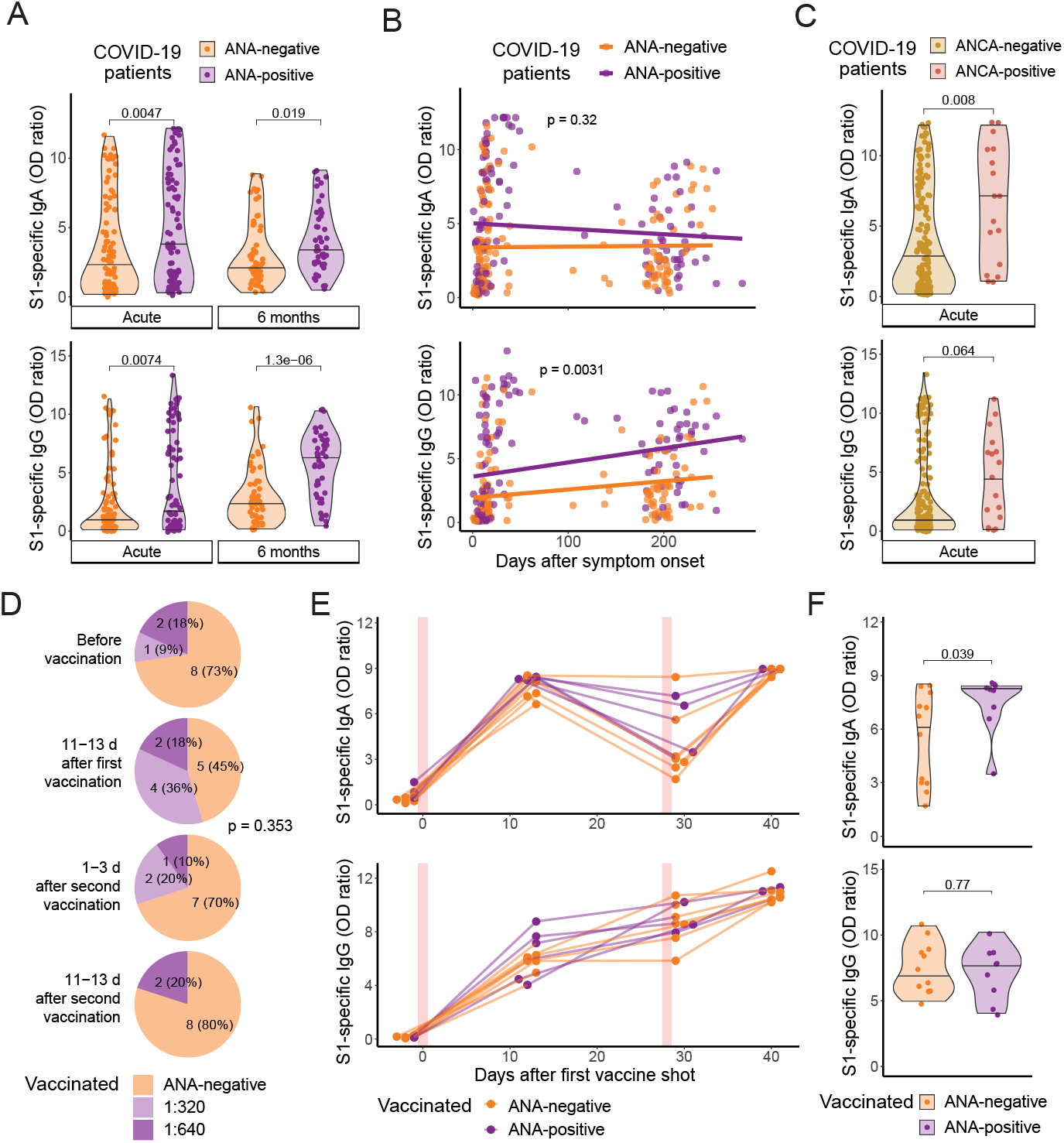
Presence of autoantibodies is associated with an increased virus-specific humoral response after SARS-CoV-2 infection and vaccination. (**A**–**B**) S1-specific IgA and IgG in ANA-positive and ANA-negative COVID-19 patients during acute disease (n = 175) and six months after recovery (n = 104). (B) P-values indicate significance of the correlation of ANA positivity as an independent parameter in a multiple linear regression model accounting for age, disease severity and sampling timepoint (**Table S3**). (**C**) S1-specific IgA and IgG in ANCA-positive and ANCA-negative COVID-19 patients during acute disease (n = 175). (**D**) ANA prevalence and titers in previously unexposed individuals (n = 11) before and after vaccination with BNT162b2 at indicated timepoints. The p-value was calculated using Chi-squared test of independence. (**E**) S1-specific IgA and IgG before and after COVID19 vaccination (n = 11). Red vertical lines indicate the timepoints of first and second vaccination with BNT162b2. (**F**) S1-specific IgA and IgG in ANA-positive and ANA-negative participants following COVID-19 vaccination with BNT162b2, combining data from 10-13d after the first (n = 11) and 1-3d after the second (n = 10) vaccination.

To elucidate whether autoantibodies were associated with an increased humoral immune response only after natural SARS-CoV-2 infection or also after other antigen-specific immune responses, we measured ANA and S1-specific antibodies in 11 individuals before and after COVID-19 vaccination with BNT162b2 (**Fig. 4D**–**F, Table S2**). Although a tendency of an increased ANA prevalence following the first vaccine shot was apparent, no significant difference was observed between sampling timepoints (**Fig. 4D**). We observed higher S1- specific IgA in ANA-positive individuals when combining data from two and four weeks after the first vaccine shot, whereas no difference was observed for IgG (**Fig. 4E** and **F**). In summary, these findings suggest the presence of autoantibodies is associated with increased S1-specific humoral responses following acute COVID-19 up to six months after recovery and following SARS-CoV-2 vaccination.

### Human virome-wide serological profiling in acute COVID-19

Next, we sought to investigate qualitative aspects of antibody responses during acute COVID- 19 with respect to previous anti-viral humoral responses in ANA-positive and ANA-negative individuals. Based on the phage immunoprecipitation sequencing (PhIP-seq) technology (VirScan),^26^ we performed human virome-wide serological profiling in 97 acute COVID-19 patients and 18 healthy controls. We assessed the results of antibodies directed to 112 different viruses (**Table S4**), with data for a total of 87,890 epitopes, consisting of 56-amino acid (AA)- long, overlapping peptides. The library comprised all six human coronaviruses (HCoV) described before the COVID-19 pandemic, including HCoV-HKU1, HCoV-NL63, HCoV- 229E, betacoronavirus 1 (BCoV1, including HCoV-OC43), severe acute respiratory syndrome-related coronavirus (SARS-CoV), and Middle east respiratory syndrome-related coronavirus (MERS-CoV).

A multivariate analysis using the summed epitope hits per viral species revealed distinct differences in COVID-19 patients compared to healthy controls, which were particularly pronounced more than one week after symptom onset (**Fig. 5A**). Between-group comparisons of COVID-19 patients and healthy controls revealed a significant difference (p < 0.005) of summed epitope hits for eight viral species (**Fig. 5B**). Of these, four enterovirus species were more abundant in healthy controls. Conversely, antibodies targeting cytomegalovirus (CMV) and Pegivirus A, and those directed to SARS-CoV and MERS-CoV were significantly more abundant in COVID-19 patients, whereas antibodies targeting the four common coronaviruses HCoV-HKU1, HCoV-NL63, HCoV-229E, and BCoV1, showed a parallel, but insignificant trend (p > 0.005) (**Fig. 5B**). Antibodies directed to all coronavirus species correlated positively with time from symptom onset (**Fig. 5C**), thus indicating production of cross-reactive antibodies during acute COVID-19.

**Figure 5.**
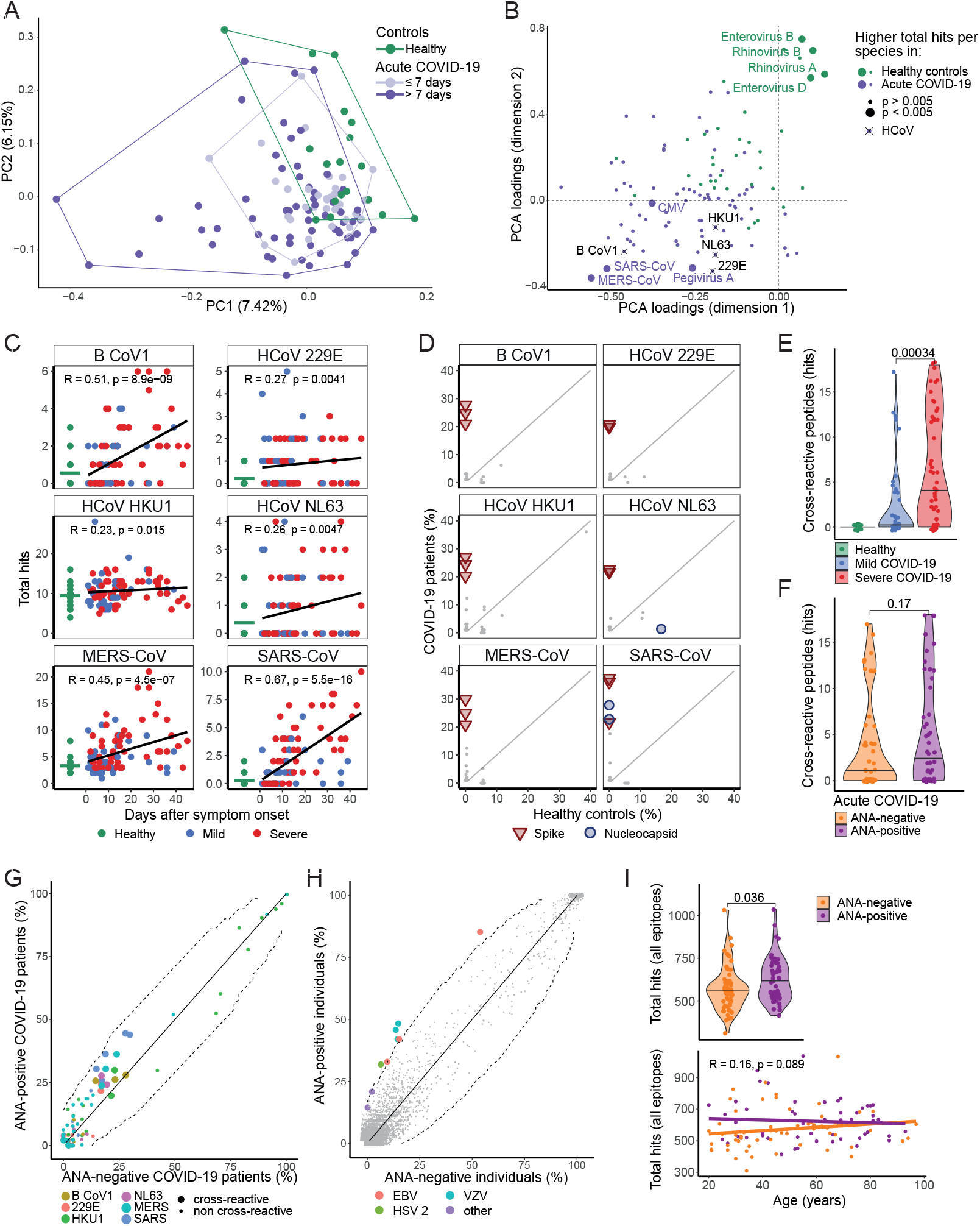
Comprehensive serological profiling (VirScan) in ANA-positive and ANA-negative individuals during acute COVID-19. (**A**) Principal component analysis (PCA) of 112 viral species, including data of 18 healthy individuals and 96 acute COVID-19 patients, grouped by timepoint of sample collection after symptom onset. Each dot represents an individual participant. (**B**) Loadings of PCA depicted in (A), with each viral species shown as individual dots (**Table S4**). Colors indicate participant groups with higher mean epitope hits per species. Viral species with significant difference (p < 0.005) between COVID-19 patients and healthy controls are shown as large colored dots. Black crosses indicate insignificant differences of coronaviruses (p > 0.005). (**C**) Temporal association of summed epitope hits of six coronaviruses after symptom onset, shown for acute COVID-19 patients (n = 97) and healthy controls (n = 18). Horizontal green bars represent means of healthy controls. (**D**) Percentage of healthy controls and COVID-19 patients with positive results for epitopes of six coronavirus species. Significantly enriched epitopes (p < 0.05) of spike and nucleocapsid are indicated accordingly. (**E**–**F**) Summed hits for cross-reactive epitopes, comparing healthy controls and patients with mild and severe COVID-19 (E) or COVID-19 patients with or without ANA (F). (**G**) Percentage of ANA-positive and ANA-negative COVID-19 patients with positive results for cross-reactive and non-cross-reactive epitopes of six coronavirus species. Dashed lines mark significance threshold at p < 0.05. (**H**) Percentage of ANA-positive and ANA-negative study participants (n = 115) with positive results, shown for all available epitopes. Significantly enriched epitopes (p < 0.005) are colored. EBV, Epstein-Barr virus; HSV-2, herpes simplex virus 2; VZV, varizella-zoster virus; other, other viruses comprising Aichivirus A and Mamastrovirus 1. (**I**) Summed epitope hits per individual including all available epitopes, comparing ANA-negative and ANA-positive participants (top; n = 115), and as a function of age (bottom).

To further study antibodies targeting CoVs in acute COVID-19, we evaluated serological profiles on a singular epitope level. We found a significantly higher (p < 0.05) proportion of COVID-19 patients tested positive for a total of 18 CoV epitopes compared to healthy controls, of which 16 were in the spike and two in the nucleoprotein (**Fig. 5D, Fig. S3A**). Since healthy individuals tested negative for these but positive for only one epitope (**Fig. 5D** and **E**), we hypothesized these 18 CoV epitopes enriched in COVID-19 patients were targeted by antibodies newly produced during acute COVID-19 and cross-reactive with shared epitopes of other CoVs. Pairwise protein alignment of these epitopes with corresponding SARS-CoV-2 proteins allowed identification of regions of SARS-CoV-2 spike and nucleoprotein targeted by cross-reactive antibodies, comprising two segments (AA positions 777–886 and 1105–1195) of spike S2 domain and one segment of nucleoprotein (AA 140–252), which have been previously identified in COVID-19 patients ^27^. Patients with severe COVID-19 tested positive for significantly more cross-reactive antibodies than mild disease patients (**Fig. 5E**). However, no significant difference was observed in ANA-positive compared to ANA-negative patients (**Fig. 5F**), although the proportion of ANA-positive patients that tested positive was slightly higher for most cross-reactive epitopes (**Fig. 5G**).

To explore potential correlations of ANA with humoral responses against other viruses, we compared seroreactivity against all tested viral epitopes and ANA positivity. Several epitopes were detected more frequently (p < 0.005) in ANA-positive, but not in ANA-negative, individuals (**Fig. 5H**). Three of the identified peptides were located on Epstein-Barr virus (EBV) nuclear antigen 2 (EBNA-2), to which ANA-positive participants showed more epitope hits, independent of age (**Fig. S3B**). When combining data of all available epitopes, we found significantly more hits in ANA-positive compared to ANA-negative participants, which was most pronounced at younger age (**Fig. 5I**). Thus, human virome-wide serological profiling in acute COVID-19 revealed antibodies cross-reactive to other coronaviruses, whereas ANA-positive participants producing antibodies to more viral epitopes on a virome wide level.

### Association of autoantibodies with inflammatory signature during acute COVID-19

Several studies have associated autoantibodies with severe COVID-19.^11,15,17-19^ Thus, we sought to further characterize ANA-positive and ANA-negative COVID-19 patients during acute disease by proteomics comprising 86 inflammatory markers, cytokine measurements, flow cytometry, clinical history, and routine diagnostic analyses. The proportion of participants with a known autoimmune disease was low in our cohort and indifferent in individuals with or without ANA or ANCA (**Table 2**). We found a higher prevalence of comorbidities in autoantibody-positive patients, including hypertension and heart disease, but no significant sex difference (**Table 2**).

**Table 2.**
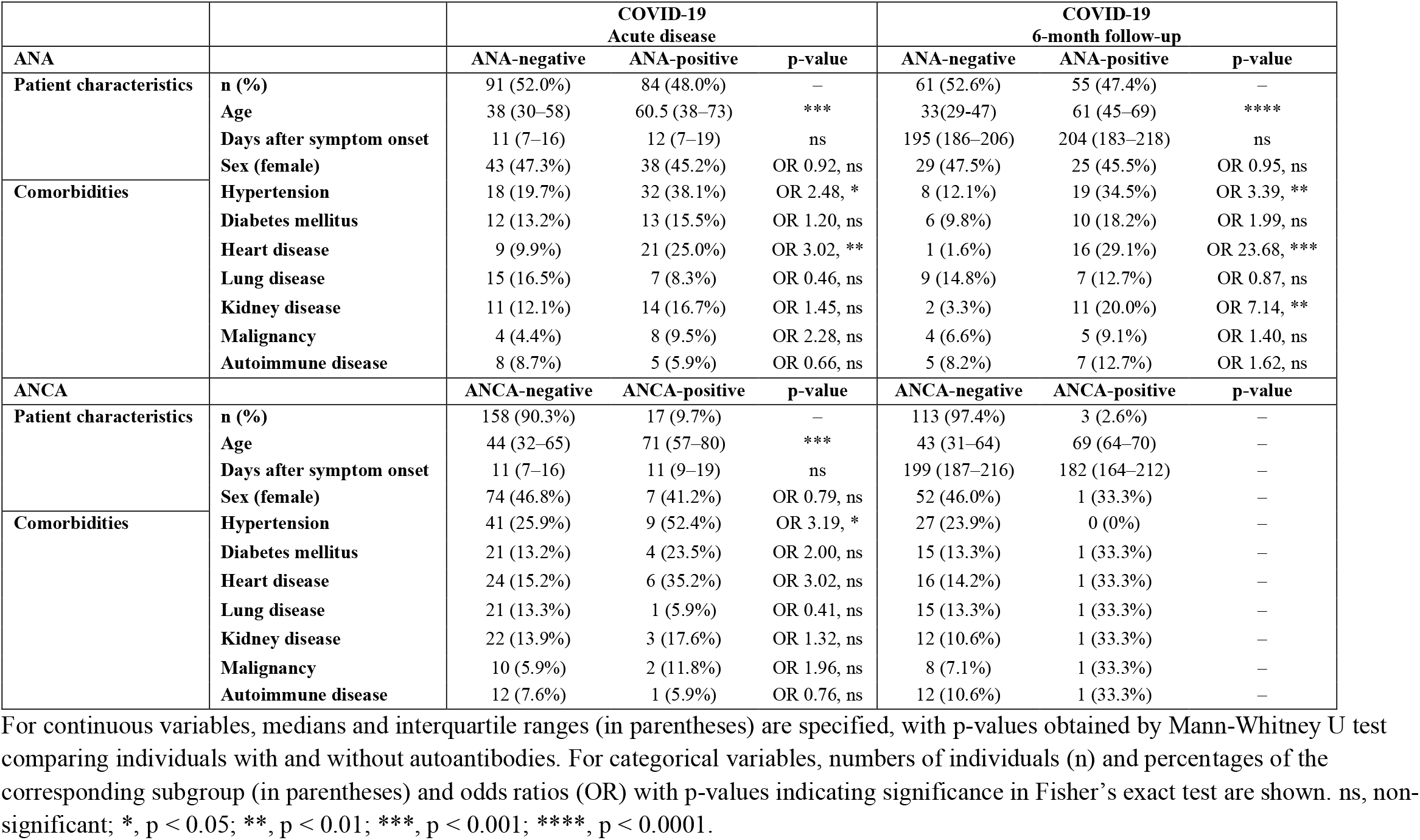
Characteristics of ANA- or ANCA-positive and -negative COVID-19 patients at acute disease or six months after infection.

A multivariate analysis with 130 variables, including demographic parameters, routine diagnostic measurements, and inflammation markers obtained by proteomics (**Table S5**), allowed for a nearly complete separation of severe COVID-19 patients from healthy individuals, with mild COVID-19 patients exhibiting intermediate characteristics (**Fig. 6A**). Several markers contributing to severe COVID-19 were significantly higher (p < 0.05) in ANA-positive than ANA-negative COVID-19 patients, revealing an inflammatory signature associated with severe disease in ANA-positive individuals (**Fig. 6B**). Importantly, ANA-positive COVID-19 patients were older and experienced longer hospitalization (**Fig. 6C**), and many inflammation markers, including C-reactive protein (CRP) and interleukin (IL)-6, were elevated compared to ANA-negative patients (**Fig. 6D**). Furthermore, ANA-positivity was associated with T cell activation as suggested by higher soluble IL-2 receptor alpha (sIL-2Rα) serum concentrations and increased proportions of activated CD38^+^ HLA-DR^+^ CD4^+^ and CD8^+^ T cells (**Fig. 6E**). Similar trends toward an inflammatory signature were also observed in ANCA-positive individuals during acute COVID-19, although these results were limited due to lower prevalence of ANCA (**Fig. S4A** and **B**).

**Figure 6.**
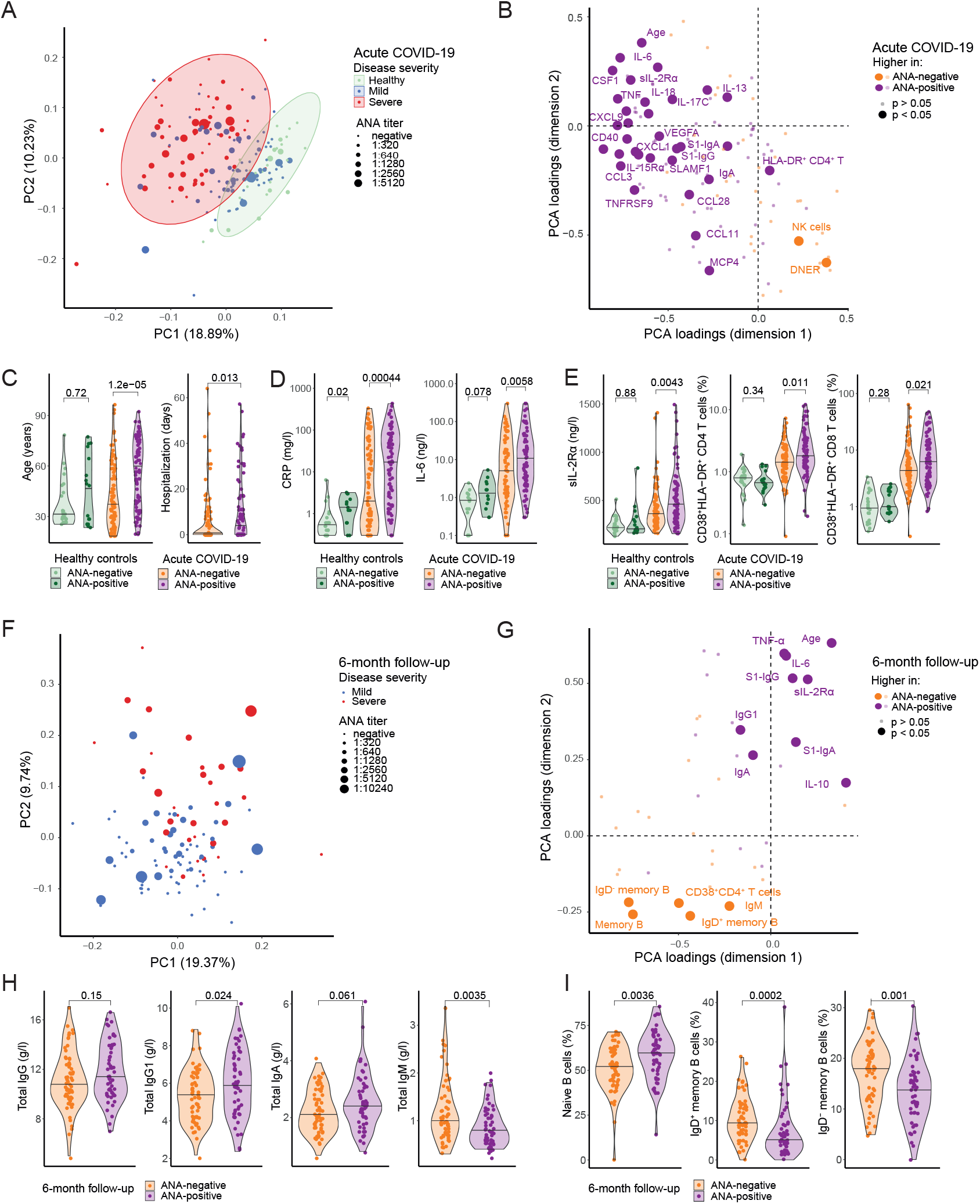
ANA-positive COVID-19 patients exhibit a pro-inflammatory signature. **(A)** PCA accounting for 130 parameters (**Table S5**) including data of healthy controls (n = 28) and acute COVID-19 patients (n = 146). Participants with missing values were excluded from this analysis. 95% confidence ellipses (t-distributed) are shown for healthy controls and severe COVID-19 patients. (**B**) Loadings (variable coordinates) of the PCA depicted in (A), with each parameter shown as an individual dot. Colors indicate the group of COVID-19 patients with higher mean for each parameter, and parameters with significant differences (p < 0.05) are represented as large dots and selected parameters are annotated (**Table S5**). (**C–E**) Comparison of ANA-negative and -positive individuals among healthy controls or acute COVID-19 patients. (C) Patient characteristics, including duration of hospitalization (n = 174) and age (n = 216). (D) Inflammation markers, including CRP (n = 209) and IL-6 (n = 215). (E) T cell activation, including sIL-2Rα (n = 215), and CD38^+^HLA-DR^+^ CD4^+^ (n = 210) and CD8^+^ (n = 209) T cells. (**F–G**) PCA (F) and loadings (G) accounting for 43 parameters (**Table S6**) including data of COVID-19 patients six months after recovery (n = 107). Participants with missing values were excluded from this analysis. (**H–I**) Comparison of ANA-negative and ANA-positive COVID-19 patients six months after recovery. (H) Concentration of total Ig subclasses in serum (n = 116). (I) Frequency of B cell subsets, including IgD^+^CD27^-^ naïve, IgD^+^CD27^+^ non-switched memory and IgD^-^CD27^+^ switched memory B cells (n = 114).

Finally, we assessed characteristics of ANA-positive and ANA-negative COVID-19 patients at six months after acute disease to identify alterations in the absence of acute inflammation. A multivariate analysis of 43 parameters, including patient characteristics and routine diagnostic measurements, revealed differences comparing ANA-positive and ANA-negative individuals, with several inflammation markers, including IL-6, tumor necrosis factor alpha (TNF-α), and sIL-2Rα, being significantly higher in ANA-positive participants (**Fig. 6F** and **G, Table S6**). Interestingly, we also observed differences in Ig subclasses, with significantly higher IgG1 and significantly lower IgM in ANA-positive individuals (**Fig. 6H**). Furthermore, marked changes in B cell subsets were apparent, with higher frequencies of IgD^+^ CD27^−^ naïve B cells and lower frequencies of IgD^+^ CD27^+^ non-switched and IgD^−^ CD27^+^ switched memory B cells in ANA-positive individuals (**Fig. 6I**). Altogether, in autoantibody-positive COVID-19 patients, we found an inflammatory signature during acute disease resembling alterations found in severe disease and changes in inflammation markers, Ig subclasses, and B cells at six months after recovery.

## Discussion

Although autoantibodies targeting nuclear, cytoplasmic, and soluble autoantigens following viral infections have been well described,^28,29^ their significance has remained ill-defined. In this study, we used highly sensitive assays to detect ANA and ANCA, representing systemic autoantibodies, in patients up to one year after infection with SARS-CoV-2. Firstly, we found transient ANA and ANCA in a subgroup of participants during acute COVID-19. Autoantibody production could result from activation of autoreactive B and T cells recognizing viral epitopes by means of molecular mimicry.^30,31^ Alternatively, antigen-independent ‘ bystander’ activation of autoreactive B and T cells by cytokines and other inflammatory mediators could drive autoantibody production.^32,33^ Particularly in severe COVID-19, which is associated with early neutrophilia and pronounced neutrophil extracellular trap (NET) formation,^34,35^ NETs expose shielded intracellular self-antigens,^25,36,37^ thus causing production of ANCA and ANA. Interestingly, a high prevalence of IgA ANCA has been reported in acute COVID-19 patients showing chilblain-like lesions.^21^ In our study, all ANCA-positive subjects tested negative for anti-MPO and anti-PR3 antibodies, thus it remains elusive whether these ANCA have pathogenic potential.

Secondly, we found distinct features in autoantibody-positive COVID-19 patients during acute disease and recovery. Autoantibodies were associated with prolonged hospitalization and inflammation markers during acute disease, supporting recent findings.^11,15,17-19,38,39^ Whereas autoantibodies targeting type I interferons have been linked to severe COVID-19,^10-12,16^ severe COVID-19 could decrease self-tolerance by tissue damage and inflammation, altogether leading to generation of autoantibodies. However, confounding factors should be considered, such as age and comorbidities, affecting prevalence of autoantibodies^40^ and risk of severe COVID-19^1^. Following these considerations, we found changes in ANA-positive individuals even six months after acute COVID-19, indicating ongoing low-grade inflammation. Furthermore, we observed alterations of the B cell compartment, including increased naïve and decreased memory B cells, previously associated with pre-symptomatic and early-stage autoimmune diseases.^41-43^ Differences in total Ig concentrations have been found in autoimmune diseases^44^ and patients suffering from post-acute COVID-19 syndrome (PACS)^45^. Thus, we have previously identified an Ig signature in PACS, including low total IgM, and found clinical risk factors, including increased age and severe disease course.^45^ Although a direct link to autoantibody development in PACS has not been reported, a misdirected immune response may underly both manifestations.

Thirdly, we observed higher S1-specific antibody titers in autoantibody-positive COVID-19 patients. Similarly, recent reports found increased anti-viral humoral responses in autoantibody-positive individuals during acute COVID-19, although the interrelation remained unclear.^16,19,22^ Interestingly, following COVID-19 mRNA vaccination in systemic lupus erythematosus (SLE) patients, higher humoral responses positively correlated with anti- dsDNA antibodies,^39^ supporting our findings of increased S1-specific IgA production in ANA- positive individuals following vaccination. These findings indicate an inherent capacity of ANA-positive individuals to mount more robust antibody responses upon antigen challenge. Human virome-wide serological profiling revealed production of cross-reactive antibodies to other coronaviruses during acute COVID-19, particularly in severe disease, consistent with broader humoral immune responses in severe COVID-19.^27^ Whereas antibodies targeted similar cross-reactive coronavirus epitopes in ANA-positive and ANA-negative COVID-19 patients, more antibodies targeted EBV antigen EBNA-2 in ANA-positive individuals. Higher humoral responses against EBV have been described in ANA-positive individuals, irrespective of autoimmune disease.^46,47^ Also, EBV has been associated with development of ANA and SLE.^48^ Furthermore, severe acute COVID-19 is characterized by extrafollicular B cell activation,^20,38,49^ which is found in autoimmune disease and associated with activation of autoreactive B cells. This increased response could allow for rapid formation of virus-specific antibody-secreting cells,^20^ potentially explaining why individuals with autoantibodies exhibit higher humoral responses during acute COVID-19. Whether autoantibody-positive subjects also show increased SARS-CoV-2-specific long-lived T cells responses^50^ remains to be investigated.

Limitations of this study include the use of highly sensitive IIF assays that yielded a high prevalence of positive results in healthy subjects and COVID-19 patients. Most of the measured ANA titers were at or just above the threshold level, which usually would be considered of irrelevant clinical significance. Moreover, we did not assess the specificity of autoantibodies, but recent studies have shown reactivity to a wide spectrum of autoantigens.^11,16^Altogether, our study shows autoantibodies in COVID-19 appear to be transient and correlate with increased anti-viral humoral immune responses and a distinct immune signature. As questions arise regarding long-term consequences of COVID-19, including the risk of immune dysregulation and autoimmune disease, understanding the mechanisms involved in balancing self-tolerance and protective immune responses become crucial to recognize and manage patients at risk for developing autoimmune diseases.

## Methods

### Human subjects and patient characteristics

Following written informed consent, adult individuals were recruited for medical history and blood sampling between April 2020 and May 2021. The study was approved by the Cantonal Ethics Committee of Zurich (BASEC #2016-01440). The cohort comprised mild and severe COVID-19 patients, healthy controls, and vaccinated individuals.

*COVID-19 patients* (**Table 1, Table S1**): 175 patients with reverse transcriptase quantitative polymerase chain reaction (RT-qPCR)-confirmed SARS-CoV-2 infection were included during acute COVID-19 at four hospitals in the Canton of Zurich, Switzerland. COVID-19 was classified for maximum disease severity according to the World Health Organization (WHO) classification criteria into mild disease – including asymptomatic (n = 4), mild illness (n = 93) and mild pneumonia (n = 12) – and severe disease – including severe pneumonia (n = 30) and acute respiratory distress syndrome (n = 36) ^51^. Follow-up visits for medical history and blood collection were conducted approximately six months and one year after symptom onset. Unreachable individuals or those declining further participation were lost to follow-up.

#### *Healthy controls* (**Table 1**)

41 participants with negative history of SARS-CoV-2 infection and serology were recruited. Five individuals developed COVID-19 after inclusion and were subsequently allocated to the patient cohort.

#### *Vaccinated individuals* (**Table S2**)

11 individuals with a negative history and serology for SARS-CoV-2 infection were sampled once before vaccination, once after the first and twice after the second mRNA vaccination with BNT162b2 (BioNTech-Pfizer).

### Autoantibody detection

ANA were measured by IIF on HEp-2 cells (Euroimmun) with a cut-off dilution of 1:320. ANCA were measured by IIF on neutrophils fixed by ethanol and formalin (Euroimmun) with a cut-off dilution of 1:40. IIF imaging was performed using a diagnostic, computer-aided microscopy system (Euroimmun). ANA patterns were classified according to the international consensus on ANA patterns anti-cell (AC) nomenclature^23^ by blinded trained personnel. For paired analyses of ANA patterns, 129 pairs of IIF pictures at 1:320 dilution were blinded for patient characteristics and sampling timepoint, and examined pairwise by the same observer. Antibodies against myeloperoxidase and proteinase 3 were measured on Phadia™ 250 (ThermoFisher Scientific) or on Bioflash^®^ (Werfen) according to manufacturer’ s instructions.

### Immunoassays

Immunoassays for Ig subsets, anti-SARS-CoV-2 spike S1-specific IgA and IgG, interleukin (IL)-1β, IL-2, IL-5, IL-6, IL-10, IL-12, interferon-γ (IFN-γ), sIL-2Rα, and tumor necrosis factor α (TNF-α), were performed in accredited laboratories at University Hospital Zurich. Serum Ig subsets were quantified on an Optilite^®^ turbidimeter (The Binding Site Group). S1- specific IgA and IgG were measured by enzyme-linked immunosorbent assays (ELISA) (Euroimmun), as established ^52^. IL-1β, IL-2, IL-6, IFN-γ and sIL-2Rα were determined by ELISA (R&D Systems) on Opsys Reader™ (Dynex). IL-5, IL-10 and IL-12 were measured by cytometric bead assays (BD Biosciences) on a Navios cytometer (Beckman Coulter). TNF-α was determined with a kit (R&D Systems) using MagPix^®^ (ThermoFisher Scientific).

### Flow cytometry

As established ^53^, blood samples were processed and analyzed in accredited laboratories at University Hospital Zurich. Blood samples were lysed with VersaLyse, fixed with IOTest3 solution and stained with antibodies (Beckman Coulter; **Table S7**). Absolute cell counts were determined using Flow Set Pro Fluorospheres calibration beads on Navios (Beckman Coulter).

### Serum proteomics

Serum samples were analyzed by a proximity extension assay-based technology 92-plex inflammation panel (Olink^®^), as established ^33,53,54^. Six parameters were excluded because less than 50% of samples showed results above detection limit.

### Human virome-wide serological profiling

As established,^7,55^ serum samples were inactivated, normalized for total IgG concentration, and incubated as duplicates with a bacteriophage library displaying linear, 56 amino acid long viral epitopes. IgG-phage complexes were captured with magnetic beads, lysed and quantified by next-generation sequencing. Blank beads samples were used as negative controls. Reads were mapped to the epitope library with Bowtie2, and counts were obtained using SAMtools. A previously described binning strategy was used to identify positivity for epitopes,^56^ with a minimum z-score of 3.5 for both sample replicates compared to negative controls. Results for a total of 112 different human viruses were included in the further analysis. (**Table S4**), whereas eukaryotes, prokaryotes, non-human viruses and human viruses with no variance or a maximal summed epitope hit count below three were excluded.

### Statistics

Statistical analyses were performed using R (version 4.1.0). Unless otherwise specified, between-group comparison was performed using two-tailed, non-parametric, unpaired testing (Mann-Whitney U) for numeric variables and odds ratios with Fisher’ s exact test for categorical variables, with p-values of <0.05 defined as significant. Missing values were omitted. Principal component analyses (PCA) were performed using stats (4.2.0) and factoextra (1.0.7) with scaled, centered variables, and loadings are shown as variable coordinates. Spearman’ s rank correlation was used for associations of numeric variables. Pairwise protein alignment for 56- amino acid (AA) long peptides with SARS-CoV-2 spike (Uniprot Entry P0DTC2) and nucleoprotein (P0DTC9) was generated using Biostrings (2.60.2), with BLOSUM62 substitution matrix and gap opening and extension penalty of -11 and -1, respectively. Data visualization was performed using ggplot2 (version 3.3.5), ggfortify (0.4.12), ggVennDiagram (1.1.4), UpSetR (1.4.0), and corrplot (0.90). Horizontal lines in violin plots represent medians. Regression lines represent simple linear regression models.

## Data Availability

All data produced in the present study are available upon reasonable request to the authors

## Acknowledgements

This work was funded by Swiss National Science Foundation grants 310030-172978 and 310030-200669 (to OB), 4078P0-198431 (to OB and JN) and NRP78 Implementation Programme (to CC and OB), Digitalization Initiative of the Zurich Higher Education Institutions Rapid-Action Call #2021.1_RAC_ID_34 (to CC), Swiss Academy of Medical Sciences grants 323530-191220 (to CC), 323530-191230 (to YZ) and 323530-177975 (to SA), Forschungskredit Candoc of University of Zurich FK-20-022 (to SA), Young Talents in Clinical Research Project Grant (YTCR 08/20) by the Swiss Academy of Medical Sciences and Bangerter Foundation (to MER), the Clinical Research Priority Program of University of Zurich for CRPP CYTIMM-Z (to OB), the Pandemic Fund of University of Zurich (to OB), and an Innovation Grant of University Hospital Zurich (to OB). We thank the diagnostic laboratories of the University Hospital Zurich, Alessandra Guaita, Claudia Meloni, Jennifer Jörger, Jana Epprecht and Claudia Bachmann for their support, and the members of the Boyman Laboratory for helpful discussions. The study overview graphic was generated with BioRender.com.

## Author contributions

P.T. contributed to patient recruitment and data collection, analysis and interpretation. C.C. contributed to patient recruitment, data collection and data interpretation. Y.Z., S.H., and S.A. contributed to patient recruitment and data collection. C.P., Z.T., and P.B. contributed to data collection. M.E.R. contributed to patient recruitment and clinical management. E.B., A.R., M.S.-H., L.C.H., and J.N. contributed to patient recruitment. E.P.-M. contributed to data analysis. O.B. conceived the project and interpreted the data. P.T. and O.B. wrote the manuscript. All authors edited and approved the final draft of the article.

## References

1. Guan WJ, Ni ZY, Hu Y, et al. Clinical Characteristics of Coronavirus Disease 2019 in China. N Engl J Med. 2020;382(18):1708–1720.

2. Petersen E, Koopmans M, Go U, et al. Comparing SARS-CoV-2 with SARS-CoV and influenza pandemics. Lancet Infect Dis. 2020;20(9):e238–e244.

3. Wiersinga WJ, Rhodes A, Cheng AC, Peacock SJ, Prescott HC. Pathophysiology, Transmission, Diagnosis, and Treatment of Coronavirus Disease 2019 (COVID-19): A Review. JAMA. 2020;324(8):782–793.

4. Liu Y, Sawalha AH, Lu Q. COVID-19 and autoimmune diseases. Curr Opin Rheumatol. 2021;33(2):155–162.

5. Zhang Y, Xiao M, Zhang S, et al. Coagulopathy and Antiphospholipid Antibodies in Patients with Covid-19. N Engl J Med. 2020;382(17):e38.

6. Toscano G, Palmerini F, Ravaglia S, et al. Guillain-Barré Syndrome Associated with SARS-CoV-2. N Engl J Med. 2020;382(26):2574–2576.

7. Consiglio CR, Cotugno N, Sardh F, et al. The Immunology of Multisystem Inflammatory Syndrome in Children with COVID-19. Cell. 2020;183(4):968–981.e967.

8. Zuo Y, Estes SK, Ali RA, et al. Prothrombotic autoantibodies in serum from patients hospitalized with COVID-19. Sci Transl Med. 2020;12(570).

9. Hsu TY, D’Silva KM, Patel NJ, Fu X, Wallace ZS, Sparks JA. Incident systemic rheumatic disease following COVID-19. Lancet Rheumatol. 2021;3(6):e402–e404.

10. Bastard P, Rosen LB, Zhang Q, et al. Autoantibodies against type I IFNs in patients with life-threatening COVID-19. Science. 2020;370(6515).

11. Wang EY, Mao T, Klein J, et al. Diverse functional autoantibodies in patients with COVID-19. Nature. 2021;595(7866):283–288.

12. Bastard P, Gervais A, Le Voyer T, et al. Autoantibodies neutralizing type I IFNs are present in. Sci Immunol. 2021;6(62).

13. Fujinami RS, von Herrath MG, Christen U, Whitton JL. Molecular mimicry, bystander activation, or viral persistence: infections and autoimmune disease. Clin Microbiol Rev. 2006;19(1):80–94.

14. Vlachoyiannopoulos PG, Magira E, Alexopoulos H, et al. Autoantibodies related to systemic autoimmune rheumatic diseases in severely ill patients with COVID-19. Ann Rheum Dis. 2020;79(12):1661–1663.

15. Sacchi MC, Tamiazzo S, Stobbione P, et al. SARS-CoV-2 infection as a trigger of autoimmune response. Clin Transl Sci. 2021;14(3):898–907.

16. Chang SE, Feng A, Meng W, et al. New-onset IgG autoantibodies in hospitalized patients with COVID-19. Nat Commun. 2021;12(1):5417.

17. Pascolini S, Vannini A, Deleonardi G, et al. COVID-19 and Immunological Dysregulation: Can Autoantibodies be Useful? Clin Transl Sci. 2021;14(2):502–508.

18. Chang SH, Minn D, Kim YK. Autoantibodies in moderate and critical cases of COVID-19 Clin Transl Sci. 2021.

19. Lerma LA, Chaudhary A, Bryan A, Morishima C, Wener MH, Fink SL. Prevalence of autoantibody responses in acute coronavirus disease 2019 (COVID-19). J Transl Autoimmun. 2020;3:100073.

20. Woodruff MC, Ramonell RP, Saini AS, et al. Relaxed peripheral tolerance drives broad. medRxiv. 2021.

21. Frumholtz L, Bouaziz JD, Battistella M, et al. Type I interferon response and vascular alteration in chilblain-like lesions during the COVID-19 outbreak. Br J Dermatol. 2021.

22. Emmenegger M, Kumar SS, Emmenegger V, et al. Anti-prothrombin autoantibodies enriched after infection with SARS-CoV-2 and influenced by strength of antibody response against SARS-CoV-2 proteins. PLoS Pathog. 2021;17(12):e1010118.

23. Chan EK, Damoiseaux J, Carballo OG, et al. Report of the First International Consensus on Standardized Nomenclature of Antinuclear Antibody HEp-2 Cell Patterns 2014-2015. Front Immunol. 2015;6:412.

24. Suwanchote S, Rachayon M, Rodsaward P, et al. Anti-neutrophil cytoplasmic antibodies and their clinical significance. Clin Rheumatol. 2018;37(4):875–884.

25. Nakazawa D, Kumar S, Desai J, Anders HJ. Neutrophil extracellular traps in tissue pathology. Histol Histopathol. 2017;32(3):203–213.

26. Xu GJ, Kula T, Xu Q, et al. Viral immunology. Comprehensive serological profiling of human populations using a synthetic human virome. Science. 2015;348(6239):aaa0698.

27. Shrock E, Fujimura E, Kula T, et al. Viral epitope profiling of COVID-19 patients reveals cross-reactivity and correlates of severity. Science. 2020;370(6520).

28. Hansen KE, Arnason J, Bridges AJ. Autoantibodies and common viral illnesses. Semin Arthritis Rheum. 1998;27(5):263–271.

29. Spohn G, Arenas-Ramirez N, Bouchaud G, Boyman O. Endogenous polyclonal anti-IL-1 antibody responses potentiate IL-1 activity during pathogenic inflammation. J Allergy Clin Immunol. 2017;139(6):1957–1965.e1953.

30. Moody R, Wilson KL, Boer JC, et al. Predicted B Cell Epitopes Highlight the Potential for COVID-19 to Drive Self-Reactive Immunity. Frontiers in Bioinformatics. 2021;1(31).

31. Angileri F, Legare S, Marino Gammazza A, Conway de Macario E, Jl Macario A, Cappello F. Molecular mimicry may explain multi-organ damage in COVID-19. Autoimmun Rev. 2020;19(8):102591.

32. Fajgenbaum DC, June CH. Cytokine Storm. N Engl J Med. 2020;383(23):2255–2273.

33. Chevrier S, Zurbuchen Y, Cervia C, et al. A distinct innate immune signature marks progression from mild to severe COVID-19. Cell Rep Med. 2021;2(1):100166.

34. Zuo Y, Yalavarthi S, Shi H, et al. Neutrophil extracellular traps in COVID-19. JCI Insight. 2020;5(11).

35. Reusch N, De Domenico E, Bonaguro L, et al. Neutrophils in COVID-19. Front Immunol. 2021;12:652470.

36. van der Linden M, van den Hoogen LL, Westerlaken GHA, et al. Neutrophil extracellular trap release is associated with antinuclear antibodies in systemic lupus erythematosus and anti-phospholipid syndrome. Rheumatology (Oxford). 2018;57(7):1228–1234.

37. Egholm C, Heeb LEM, Impellizzieri D, Boyman O. The Regulatory Effects of Interleukin-4 Receptor Signaling on Neutrophils in Type 2 Immune Responses. Front Immunol. 2019;10:2507.

38. Woodruff MC, Ramonell RP, Nguyen DC, et al. Extrafollicular B cell responses correlate with neutralizing antibodies and morbidity in COVID-19. Nat Immunol. 2020;21(12):1506–1516.

39. Izmirly PM, Kim MY, Samanovic M, et al. Evaluation of Immune Response and Disease Status in SLE Patients Following SARS-CoV-2 Vaccination. Arthritis Rheumatol. 2021.

40. Dinse GE, Parks CG, Weinberg CR, et al. Increasing Prevalence of Antinuclear Antibodies in the United States. Arthritis Rheumatol. 2020;72(6):1026–1035.

41. Baglaenko Y, Chang NH, Johnson SR, et al. The presence of anti-nuclear antibodies alone is associated with changes in B cell activation and T follicular helper cells similar to those in systemic autoimmune rheumatic disease. Arthritis Res Ther. 2018;20(1):264.

42. Zhu L, Yin Z, Ju B, et al. Altered frequencies of memory B cells in new-onset systemic lupus erythematosus patients. Clin Rheumatol. 2018;37(1):205–212.

43. Slight-Webb S, Lu R, Ritterhouse LL, et al. Autoantibody-Positive Healthy Individuals Display Unique Immune Profiles That May Regulate Autoimmunity. Arthritis Rheumatol. 2016;68(10):2492–2502.

44. Zhang H, Li P, Wu D, et al. Serum IgG subclasses in autoimmune diseases. Medicine (Baltimore). 2015;94(2):e387.

45. Cervia C, Zurbuchen Y, Taeschler P, et al. Immunoglobulin signature predicts risk of post-acute COVID-19 syndrome. Nat Commun. 2022;13(1):446.

46. Slight-Webb S, Smith M, Bylinska A, et al. Autoantibody-positive healthy individuals with lower lupus risk display a unique immune endotype. J Allergy Clin Immunol. 2020;146(6):1419–1433.

47. Jog NR, Young KA, Munroe ME, et al. Association of Epstein-Barr virus serological reactivation with transitioning to systemic lupus erythematosus in at-risk individuals. Ann Rheum Dis. 2019;78(9):1235–1241.

48. Cuomo L, Cirone M, Di Gregorio AO, et al. Elevated antinuclear antibodies and altered anti-Epstein-Barr virus immune responses. Virus Res. 2015;195:95–99.

49. Kaneko N, Kuo HH, Boucau J, et al. Loss of Bcl-6-Expressing T Follicular Helper Cells and Germinal Centers in COVID-19. Cell. 2020;183(1):143–157.e113.

50. Adamo S, Michler J, Zurbuchen Y, et al. Signature of long-lived memory CD8. Nature. 2021.

51. WHO. COVID-19 Clinical management: living guidance. World Health Organization (2021), doi: www.who.int/publications/i/item/WHO-2019-nCoV-clinical-2021-1. In.

52. Cervia C, Nilsson J, Zurbuchen Y, et al. Systemic and mucosal antibody responses specific to SARS-CoV-2 during mild versus severe COVID-19. J Allergy Clin Immunol. 2021;147(2):545–557.e549.

53. Adamo S, Chevrier S, Cervia C, et al. Profound dysregulation of T cell homeostasis and function in patients with severe COVID-19. Allergy. 2021;76(9):2866–2881.

54. Lundberg M, Eriksson A, Tran B, Assarsson E, Fredriksson S. Homogeneous antibody-based proximity extension assays provide sensitive and specific detection of low-abundant proteins in human blood. Nucleic Acids Res. 2011;39(15):e102.

55. Pou C, Nkulikiyimfura D, Henckel E, et al. The repertoire of maternal anti-viral antibodies in human newborns. Nat Med. 2019;25(4):591–596.

56. Mina MJ, Kula T, Leng Y, et al. Measles virus infection diminishes preexisting antibodies that offer protection from other pathogens. Science. 2019;366(6465):599–606.

