## Supplementary for "Autoantibodies in COVID-19 correlate with anti-viral humoral responses and distinct immune signatures"

### Supplementary Figures

**Figure S1. ANA patterns in healthy controls and COVID-19 patients during acute disease and follow-up.** (**A–D**) Intersection plots showing the counts of all ANA patterns (horizontal bars) and counts of pattern combinations (vertical bars) as indicated by the dot matrix, as observed in healthy individuals (A; n = 41), acute COVID-19 patients (B; n = 175), and COVID-19 patients six months (C; n = 116) and one year (D; n = 92) after recovery.

**Figure S2. S1-specific humoral response in ANA-positive and ANA-negative COVID-19 patients one year after recovery.** S1-specific IgA and IgG in ANA-positive and ANA-negative COVID-19 patients (n = 37).

**Figure S3. Human virome-wide serological profiling in healthy controls and acute COVID-19 patients.** (**A**) Percentage of healthy controls (n = 18) and COVID-19 patients (n = 97) with positive results for all available epitopes of six coronavirus species. Significantly enriched epitopes ($p < 0.05$) are colored and shaped according to the corresponding protein. Dashed lines indicate significance threshold ($p < 0.05$). (**B**) Summed epitope hits of EBNA-2 and correlation with age in in ANA-negative and -positive participants (n = 115).

**Figure S4. Multivariate characterization of ANCA-positive and ANCA-negative acute COVID-19 patients.** (**A**) PCA accounting for 130 parameters including data of healthy controls (n = 28) and acute COVID-19 patients (n = 146). Participants with missing values were excluded from this analysis. (**B**) Loadings (variable coordinates) of the PCA depicted in (A), with each parameter shown as an individual dot (**Table S5**). Colors indicate participant group with higher mean, and parameters with significant differences ($p < 0.05$) comparing ANCA-positive and ANCA-negative COVID-19 patients are shown as large dots and annotated.

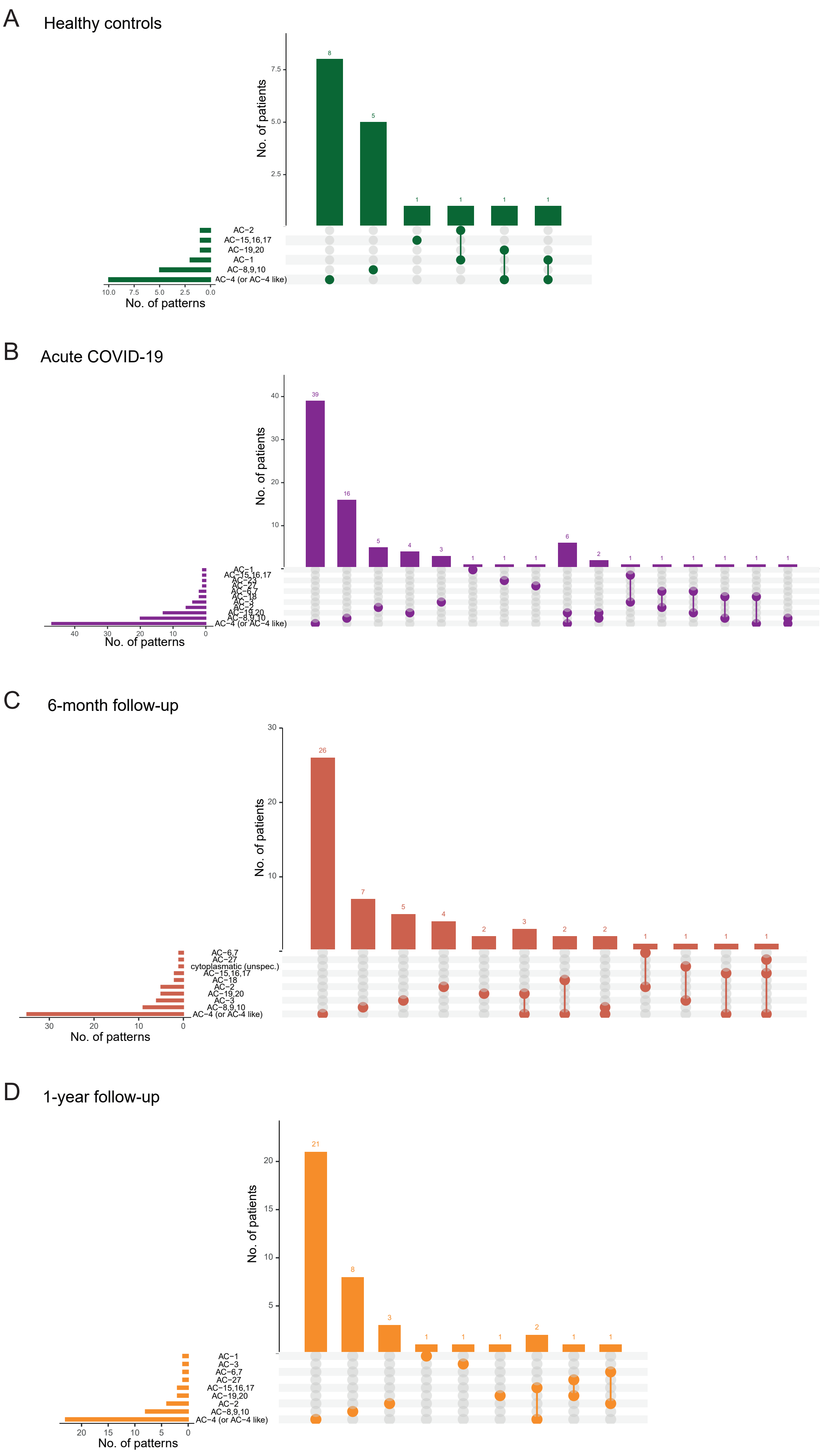


Figure S1_Taeschler et al.

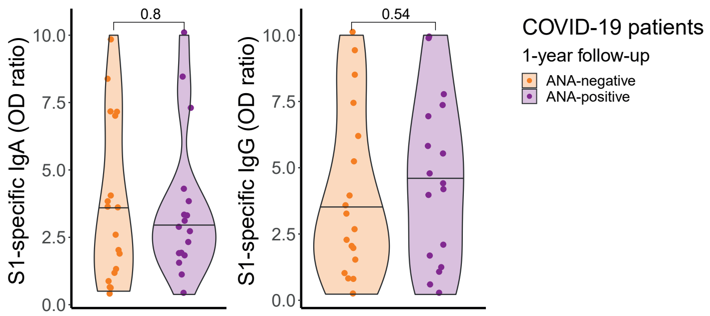


Figure S2_Taeschler et al.

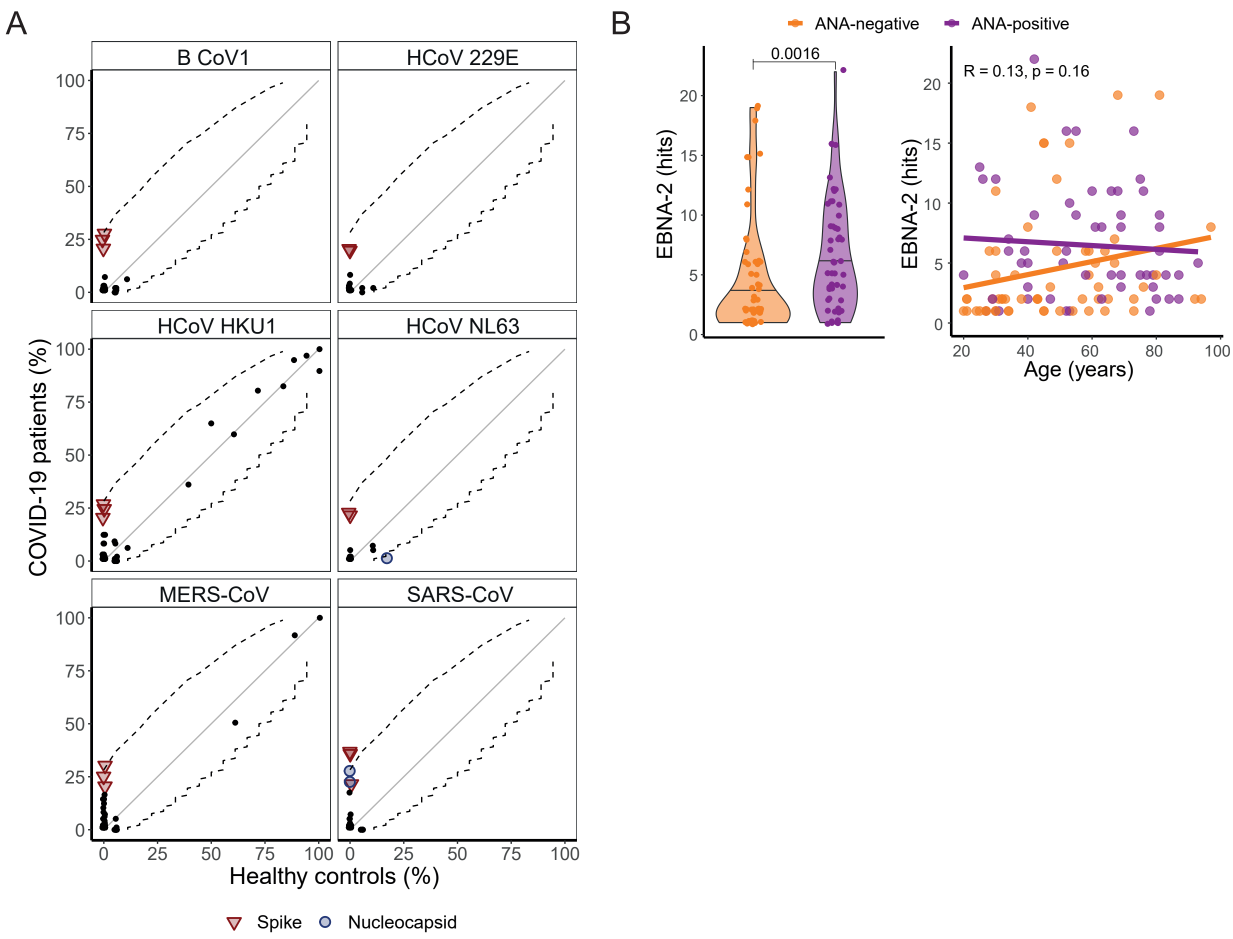


Figure S3_Taeschler et al.

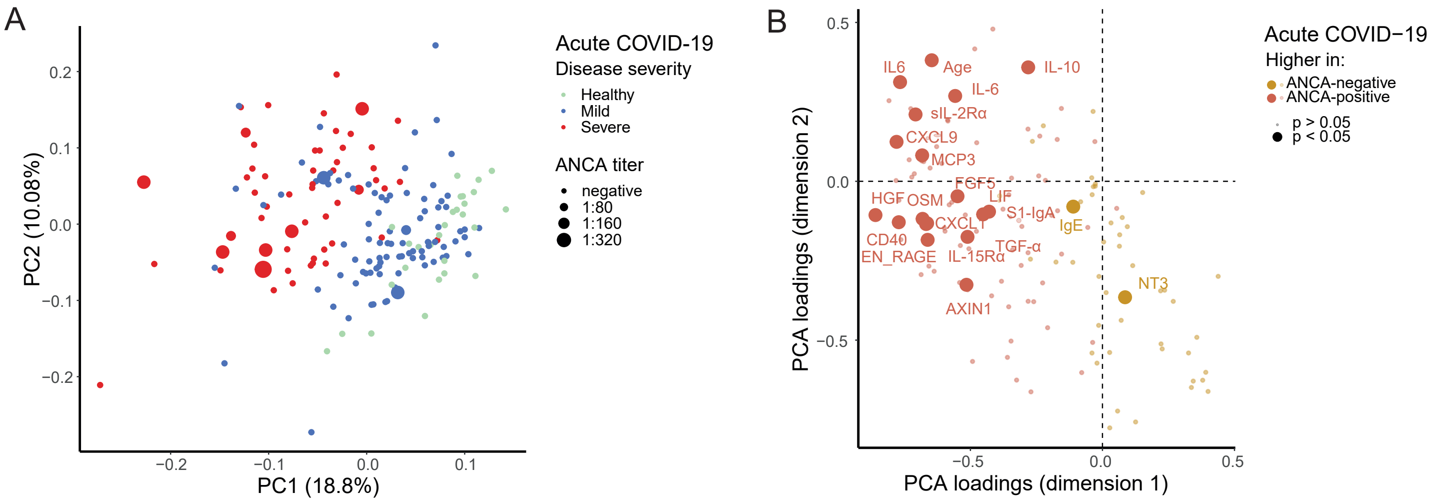


Figure S4_Taeschler et al.

| | | COVID-19 1-year follow-up | |
|---|---|---|---|
| | **Disease severity** | **Mild** | **Severe** |
| | **n (%)** | 64 (69.6%) | 28 (30.4%) |
| **Patient characteristics** | **Age** | 36 (31–52) | 56 (36–69) |
| | **Days after symptom onset** | 376 (366–387) | 380 (367–396) |
| | **Sex (female)** | 34 (53.1%) | 8 (28.6%) |
| | **Vaccinated** | 42 (65.6%) | 13 (46.4%) |
| | **Hospitalized** | 8 (12.5%) | 28 (100%) |
| **Laboratory parameters** | **Lymphocyte count (G/l)** | 1.94 (1.58–2.37) | 1.89 (1.63–2.60) |
| | **CRP (mg/l)** | 1 (0.6–2.5) | 1.6 (0.7–3.0) |
| | **TNF-α (ng/l)** | 9.2 (7.6–11.3) | 9.5 (7.5–11.6) |
| | **IL-6 (ng/l)** | 1.2 (0.2–2.1) | 1.8 (0.7–3) |
| | **S1-specific IgA (OD ratio)** | 8.38 (3.9–10.0) | 7.49 (3.24–8.59) |
| | **S1-specific IgG (OD ratio)** | 10.0 (5.60–10.0) | 9.29 (5.12–10.0) |
| **Comorbidities** | **Hypertension** | 6 (9.4%) | 15 (53.6%) |
| | **Diabetes** | 3 (4.7%) | 6 (21.4%) |
| | **Heart disease** | 2 (3.1%) | 9 (32.1%) |
| | **Lung disease** | 5 (7.8%) | 8 (28.6%) |
| | **Malignancy** | 0 | 7 (25.0%) |
| | **Kidney disease** | 3 (4.7%) | 9 (32.1%) |
| | **Autoimmune disease** | 6 (9.4%) | 3 (10.7%) |

**Table S1. Characteristics of COVID-19 patients at one year after infection.** For continuous variables, medians and interquartile ranges (in parentheses) are specified. Numbers of individuals (n) and percentages of the corresponding subgroup (in parentheses) are shown for categorical variables. OD, optical density.

| | | Vaccinated individuals without prior COVID-19 | | |
|---|---|---|---|---|
| **Blood sampling timepoint** | | **1–3 days before vaccination** | **11–13 days after first vaccination** | **1–3 days after second vaccination** |
| **Patient characteristics** | **n** | 11 | 11 | 10 |
| | **Age** | 29 (27–30) | 29 (27–30) | 29 (27–31) |
| | **Sex (female)** | 5 (45.5%) | 5 (45.5%) | 4 (40%) |
| **Laboratory parameteres** | **ANA-positive** | 3 (27.3%) | 6 (54.5%) | 3 (30%) |
| | **S1-specific IgA [OD ratio]** | 0.34 (0.3–0.5) | 8.29 (7.73–8.43)**** | 3.33 (2.87–6.31)*** |
| | **S1-specific IgG [OD ratio]** | 0.12 (0.12–0.16) | 5.92 (5.38–6.70)**** | 8.59 (8.10–9.79)*** |

**Table S2. Characteristics of vaccinated individuals before vaccination and after first and second vaccination with BNT162b2.** Medians and interquartile ranges (in brackets) are specified for continuous variables, with p-values obtained by Wilcoxon signed rank test compared to before vaccination. Numbers of individuals (n) and percentages of corresponding subgroup (in parentheses) are shown for categorical variables. ***, $p < 0.001$; ****, $p < 0.0001$; OD, optical density.

| S1 specific IgA (OD ratio, square root) | | | |
|---|---|---|---|
| **Parameter** | **Estimate** | **95% CI** | **p-value** |
| Intercept | 1.196 | 0.918–1.473 | **<0.0001** |
| Age [years] | 0.008 | 0.002–0.014 | **0.013** |
| Disease severity (*severe*) | 0.462 | 0.216–0.708 | **0.00026** |
| Sampling timepoint (*6 months*) | -0.017 | -0.208–0.174 | 0.862 |
| ANA (*positive*) | 0.122 | -0.078–0.323 | 0.231 |
| **S1 specific IgG (OD-ratio, square root)** | | | |
| **Parameter** | **Estimate** | **95% CI** | **p-value** |
| Intercept | 0.689 | 0.403–0.976 | **<0.0001** |
| Age [years] | 0.0068 | 0.0002–0.0134 | **0.048** |
| Disease severity (*severe*) | 0.595 | 0.341–0.894 | **<0.0001** |
| Sampling timepoint (*6 months*) | 0.543 | 0.345–0.740 | **<0.0001** |
| ANA (*positive*) | 0.314 | 0.107–0.522 | **0.0031** |

**Table S3. Multiple linear regression for S1-specific IgA and IgG.** Analysis accounting for age (continuous; years), disease severity (categorical; mild or severe disease), sampling timepoint (categorical; acute or 6 months) and presence of ANA (categorical; negative or positive). All observations of unvaccinated COVID-19 patients during acute COVID-19 (n = 175) and 6 months after recovery (n = 104) were included in the analysis. OD, optical density.

| **Viral species** | **Average peptide hits** | **Mean fold change COVID-19 / healthy** | **p-value** |
|---|---|---|---|
| Human herpesvirus 4 | 64.08 | 1.06 | ns |
| Human herpesvirus 5 | 61.33 | 1.53 | *** |
| Human herpesvirus 1 | 46.38 | 1.14 | ns |
| Orf virus | 24.95 | 1.09 | * |
| Human herpesvirus 2 | 24.94 | 1.22 | ns |
| Influenza A virus | 24.75 | 1.05 | ns |
| Enterovirus B | 22.43 | 0.53 | *** |
| Rhinovirus A | 20.53 | 0.47 | **** |
| Human herpesvirus 3 | 17.86 | 1.09 | ns |
| Enterovirus C | 16.96 | 0.58 | * |
| Molluscum contagiosum virus | 16.70 | 1.09 | * |
| Human respiratory syncytial virus | 11.92 | 0.81 | ns |
| Hepatitis B virus | 11.89 | 1.97 | ns |
| Hepatitis C virus | 10.91 | 1.12 | ns |
| Human coronavirus HKU1 | 10.54 | 1.14 | ns |
| Human adenovirus C | 10.11 | 0.98 | ns |
| Human adenovirus B | 9.18 | 1.10 | ns |
| Chikungunya virus | 8.45 | 1.11 | ns |
| Pegivirus A | 7.91 | 1.15 | *** |
| Rhinovirus B | 6.82 | 0.43 | **** |
| Influenza B virus | 6.33 | 0.57 | ns |
| Human herpesvirus 8 | 6.10 | 1.03 | ns |
| Rubella virus | 5.76 | 1.20 | ns |
| Human immunodeficiency virus 1 | 5.75 | 1.52 | ns |
| Hepatitis E virus | 5.73 | 1.27 | ns |
| Middle East respiratory syndrome coronavirus | 5.52 | 1.78 | *** |
| Human herpesvirus 6A | 5.04 | 1.05 | ns |
| Dengue virus | 4.84 | 2.34 | * |
| Enterovirus A | 4.33 | 0.51 | * |
| Human adenovirus D | 4.24 | 1.13 | ns |
| Alphapapillomavirus 3 | 4.19 | 0.95 | ns |
| Cowpox virus | 4.09 | 1.04 | ns |
| Human adenovirus E | 4.03 | 1.16 | ns |
| Venezuelan equine encephalitis virus | 3.65 | 1.13 | ns |
| Human herpesvirus 6B | 3.64 | 0.97 | ns |
| Ross River virus | 3.48 | 0.99 | ns |
| Betapapillomavirus 1 | 3.25 | 1.01 | ns |
| Human adenovirus F | 3.16 | 1.06 | ns |
| Sudan ebolavirus | 2.99 | 1.09 | ns |
| Zaire ebolavirus | 2.95 | 1.21 | ns |
| Alphapapillomavirus 2 | 2.70 | 1.16 | ns |
| Human adenovirus A | 2.66 | 1.13 | ns |
| Merkel cell polyomavirus | 2.65 | 0.92 | ns |
| Mamastrovirus 1 | 2.58 | 1.26 | ns |
| Alphapapillomavirus 6 | 2.17 | 0.97 | ns |
| Influenza C virus | 2.15 | 0.59 | ns |
| Alphapapillomavirus 4 | 2.05 | 1.03 | ns |
| Bundibugyo ebolavirus | 2.04 | 1.06 | ns |
| Adeno-associated dependoparvovirus A | 2.01 | 1.20 | ns |
| Sapporo virus | 1.98 | 0.82 | ns |
| Yellow fever virus | 1.96 | 1.43 | ns |
| Cosavirus A | 1.94 | 1.19 | ns |
| Torque teno virus | 1.91 | 0.95 | ns |
| Severe acute respiratory syndrome-related coronavirus | 1.90 | 7.95 | **** |
| Rotavirus A | 1.89 | 1.74 | ns |
| Variola virus | 1.87 | 1.10 | ns |
| Human herpesvirus 7 | 1.82 | 0.45 | * |
| Vaccinia virus | 1.65 | 0.99 | ns |
| Alphapapillomavirus 10 | 1.61 | 1.09 | ns |
| Colorado tick fever virus | 1.59 | 1.23 | ns |
| O'nyong-nyong virus | 1.57 | 1.34 | * |

| | | | |
|---|---|---|---|
| Eastern equine encephalitis virus | 1.56 | 0.82 | ns |
| Norwalk virus | 1.55 | 0.66 | ns |
| Alphapapillomavirus 7 | 1.48 | 1.19 | ns |
| Alphapapillomavirus 9 | 1.42 | 1.41 | ns |
| Human parvovirus B19 | 1.34 | 0.44 | ns |
| Powassan virus | 1.32 | 0.89 | ns |
| Pseudocowpox virus | 1.28 | 1.12 | ns |
| Betapapillomavirus 2 | 1.27 | 2.83 | ns |
| Louping ill virus | 1.26 | 1.10 | ns |
| Human immunodeficiency virus 2 | 1.25 | 0.93 | ns |
| Betacoronavirus 1 | 1.25 | 2.48 | * |
| Rosavirus A | 1.24 | 1.07 | ns |
| Hepatitis delta virus | 1.18 | 1.92 | ns |
| Sindbis virus | 1.18 | 1.08 | ns |
| Measles virus | 1.12 | 0.77 | ns |
| Tanapox virus | 1.04 | 1.20 | ns |
| Hepatitis A virus | 0.96 | 3.22 | ns |
| Salivirus A | 0.82 | 1.41 | ns |
| Human coronavirus 229E | 0.76 | 3.89 | * |
| Human coronavirus NL63 | 0.76 | 2.14 | ns |
| Aichivirus A | 0.67 | 1.40 | ns |
| Enterovirus D | 0.67 | 0.36 | *** |
| Yaba monkey tumor virus | 0.65 | 2.59 | ns |
| Dugbe virus | 0.60 | 1.63 | ns |
| Human parainfluenza virus 3 | 0.60 | 0.48 | ns |
| Lassa virus | 0.58 | 2.29 | ns |
| Reston ebolavirus | 0.55 | 1.29 | ns |
| Human metapneumovirus | 0.54 | 0.87 | ns |
| West Nile virus | 0.53 | 11.06 | * |
| Rabies virus | 0.51 | 0.49 | ns |
| Human parainfluenza virus 4 | 0.45 | 1.01 | ns |
| Marburg marburgvirus | 0.44 | 0.98 | ns |
| Puumala virus | 0.41 | 1.58 | ns |
| Mumps virus | 0.40 | 4.13 | ns |
| Human picobirnavirus | 0.39 | 0.87 | ns |
| Japanese encephalitis virus | 0.38 | 0.82 | ns |
| Alphapapillomavirus 5 | 0.37 | 2.44 | ns |
| Simian virus 40 (SV40) | 0.35 | 3.56 | ns |
| Rotavirus B | 0.34 | 1.28 | ns |
| WU Polyomavirus | 0.32 | 0.68 | ns |
| Human parechovirus | 0.32 | 0.22 | * |
| Vesicular stomatitis Indiana virus | 0.32 | 3.19 | ns |
| JC polyomavirus | 0.29 | 1.05 | ns |
| Alphapapillomavirus 1 | 0.29 | 1.36 | ns |
| Crimean-Congo hemorrhagic fever virus | 0.27 | 1.75 | ns |
| Monkeypox virus | 0.25 | 0.90 | ns |
| Human parainfluenza virus 2 | 0.25 | 1.63 | ns |
| California encephalitis virus | 0.19 | Inf | ns |
| Human erythrovirus V9 | 0.17 | 0.41 | ns |
| Junin virus | 0.15 | 3.00 | ns |
| Mupapillomavirus 2 | 0.15 | 0.88 | ns |

**Table S4. Viral species included in the analysis of phage immunoprecipitation sequencing (PhIP-seq)-based human virome-wide serological profiling (VirScan)**. Average of summed epitope hits per species and the ratio of mean summed hits comparing COVID-19 patients (n = 97) and healthy individuals (n = 18) are indicated. P-values were calculated using Mann-Whitney U test. ns, non-significant; *, $p < 0.05$; **, $p < 0.01$; ***, $p < 0.001$; ****, $p < 0.0001$.

| Variable name | Description | Method | Mean fold change ANA-pos / ANA-neg | p-value |
|---|---|---|---|---|
| Age | | History | 1.27 | **** |
| Days | Days after symptom onset | History | 1.22 | ns |
| IL-8 | Interleukin 8 | Olink | 1.09 | ns |
| VEGFA | Vascular epithelial growth factor A | Olink | 1.03 | *** |
| CD8A | CD8α | Olink | 1.05 | ns |
| MCP3 | Monocyte chemotactic protein 3 | Olink | 1.16 | ns |
| GDNF | Glial cell line-derived neurotrophic factor | Olink | 1.03 | * |
| CDCP1 | CUB domain-containing protein 1 | Olink | 1.34 | *** |
| CD244 | Natural killer cell receptor 2B4 | Olink | 0.98 | ns |
| IL-7 | Interleukin-7 | Olink | 1.02 | ns |
| OPG | Osteoprotegerin | Olink | 1.02 | *** |
| LAP_TGFbeta1 | Latency-associated peptide transforming growth factor β-1 | Olink | 1.03 | * |
| uPA | Urokinase-type plasminogen activator | Olink | 1.00 | ns |
| IL-6 | Interleukin-6 | Olink | 1.23 | *** |
| IL-17C | Interleukin-17C | Olink | 1.10 | * |
| MCP1 | Monocyte chemotactic protein 1 | Olink | 1.02 | ns |
| IL-17A | Interleukin-17A | Olink | 0.93 | ns |
| CXCL11 | C-X-C motif chemokine 11 | Olink | 1.04 | ns |
| AXIN1 | Axin-1 | Olink | 1.03 | ns |
| TRAIL | TNF-related apoptosis-inducing ligand | Olink | 0.99 | ns |
| IL-20Rα | IL-20 receptor subunit α | Olink | 0.96 | ns |
| CXCL9 | C-X-C motif chemokine 9 | Olink | 1.10 | **** |
| CST5 | Cystatin D | Olink | 1.02 | ns |
| IL-2Rβ | IL-2 receptor subunit β | Olink | 1.01 | ns |
| OSM | Oncostatin-M | Olink | 1.05 | ns |
| CXCL1 | C-X-C motif chemokine 1 | Olink | 1.03 | * |
| TSLP | Thymic stromal lymphoprotein | Olink | 1.09 | ns |
| CCL4 | C-C motif chemokine 4 | Olink | 1.04 | ns |
| CD6 | CD6 | Olink | 1.00 | ns |
| SCF | Stem cell factor | Olink | 0.98 | ns |
| IL-18 | Interleukin-18 | Olink | 1.03 | * |
| SLAMF1 | Signaling lymphocytic activation molecule 1 | Olink | 1.17 | **** |
| TGFalpha | Transforming growth factor α | Olink | 1.07 | ns |
| MCP4 | Monocyte chemotactic protein 4 | Olink | 1.02 | * |
| CCL11 | C-C motif chemokine 11 | Olink | 1.04 | * |
| TNFSF14 | TNF ligand superfamily member 14 | Olink | 1.02 | ns |
| FGF23 | Fibroblast growth factor 23 | Olink | 1.15 | ns |
| IL-10Rα | IL-10 receptor subunit α | Olink | 0.88 | ns |
| FGF5 | Fibroblast growth factor 5 | Olink | 1.13 | * |
| MMP1 | Matrix metalloprotease 1 | Olink | 1.03 | ns |
| LIF_R | Leukemia inhibitory factor receptor | Olink | 0.99 | ns |
| FGF21 | Fibroblast growth factor 21 | Olink | 1.07 | ns |
| CCL19 | C-C motif chemokine 19 | Olink | 1.03 | ns |
| IL-15Rα | IL-15 receptor subunit α | Olink | 1.25 | *** |
| IL-10Rβ | IL-10 receptor subunit α | Olink | 1.05 | ns |
| IL-18R1 | IL-18 receptor 1 | Olink | 1.00 | ns |
| PD_L1 | Programmed cell death 1 ligand 1 | Olink | 1.00 | ns |
| CXCL5 | C-X-C motif chemokine 5 | Olink | 1.04 | ns |
| TRANCE | TNF related activation-induced cytokine | Olink | 0.99 | ns |
| HGF | Hepatocyte growth factor | Olink | 1.09 | *** |
| IL-12β | Interleukin-12 subunit β | Olink | 1.10 | ns |
| IL-24 | Interleukin-24 | Olink | 1.21 | ns |
| IL-13 | Interleukin-13 | Olink | 1.31 | * |
| MMP10 | Matrix metalloprotease 10 | Olink | 1.02 | ns |
| IL-10 | Interleukin-10 | Olink | 1.01 | ns |
| TNF | Tumor necrosis factor | Olink | 1.08 | * |
| CCL23 | C-C motif chemokine 23 | Olink | 1.02 | ns |
| CD5 | CD5 | Olink | 1.03 | ns |
| CCL3 | C-C motif chemokine 3 | Olink | 1.08 | * |
| Flt3L | Fms-related tyrosine kinase 3 ligand | Olink | 1.00 | ns |

| | | | | |
|---|---|---|---|---|
| CXCL6 | C-X-C motif chemokine 6 | Olink | 1.02 | ns |
| CXCL10 | C-X-C motif chemokine 10 | Olink | 0.99 | ns |
| E4_BP1 | Eukaryotic translation initiation factor 4E-binding protein 1 | Olink | 1.06 | * |
| IL-20 | Interleukin-20 | Olink | 0.97 | ns |
| SIRT2 | SIR2-like protein 2 | Olink | 1.03 | ns |
| CCL28 | C-C motif chemokine 28 | Olink | 1.13 | *** |
| DNER | Delta and Notch-like epidermal growth factor-related receptor | Olink | 0.98 | * |
| EN_RAGE | Prtoein S100-A12 | Olink | 1.03 | ns |
| CD40 | CD40L receptor | Olink | 1.03 | * |
| IFNgamma | Interferon-γ | Olink | 0.94 | ns |
| FGF19 | Fibroblast growth factor 19 | Olink | 0.99 | ns |
| IL-4 | Interleukin-4 | Olink | 0.42 | ns |
| LIF | Leukemia inhibitory factor | Olink | 1.35 | * |
| NRTN | Neurturin | Olink | 0.93 | ns |
| MCP2 | Monocyte chemotactic protein 2 | Olink | 0.98 | ns |
| CASP8 | Caspase-8 | Olink | 1.00 | ns |
| CCL25 | C-C motif chemokine 25 | Olink | 1.03 | ns |
| CX3CL1 | Fractalkine | Olink | 1.02 | ns |
| TNFRSF9 | TNF receptor superfamily member 9 | Olink | 1.08 | *** |
| NT3 | Neurotrophin-3 | Olink | 1.01 | ns |
| TWEAK | TNF superfamily memebr 12 | Olink | 1.02 | ns |
| CCL20 | C-C motif chemokine 20 | Olink | 1.06 | ns |
| ST1A1 | Sulfotransferase 1A1 | Olink | 0.97 | ns |
| STAMBP | STAM-binding protein | Olink | 0.98 | ns |
| IL-5 | Interleukin-5 | Olink | 0.96 | ns |
| ADA | Adenosin deaminase | Olink | 0.97 | ns |
| TNFB | Tumor necrosis factor β | Olink | 0.98 | ns |
| CSF1 | Macrophage colony-stimulating factor 1 | Olink | 1.01 | * |
| S1-IgA | S1-specific IgA | ELISA | 2.12 | **** |
| S1-IgG | S1-specific IgG | ELISA | 2.07 | *** |
| IgG | IgG | ELISA | 1.06 | ns |
| IgG1 | IgG1 | ELISA | 1.08 | ns |
| IgG2 | IgG2 | ELISA | 0.99 | ns |
| IgG3 | IgG3 | ELISA | 1.02 | ns |
| IgG4 | IgG4 | ELISA | 0.79 | ns |
| IgM | IgM | ELISA | 0.92 | ns |
| IgA | IgA | ELISA | 1.21 | **** |
| IgD | IgD | ELISA | 1.10 | ns |
| IgE | IgE | ELISA | 1.11 | ns |
| IL-1β | Interleukin-1β | ELISA | 8.40 | ns |
| IL-2 | Interleukin-2 | ELISA | 2.27 | ns |
| sIL-2Rα | Soluble IL-2 receptor subunit α | ELISA | 1.32 | *** |
| IL-5 | Interleukin-5 | ELISA | 1.45 | ns |
| IL-6 | Interlukin-6 | ELISA | 1.87 | *** |
| IL-10 | Interluekin-10 | ELISA | 0.68 | ns |
| IL-12 | Interleukin-12 | ELISA | 0.62 | ns |
| IFN-γ | Interferon-γ | ELISA | 0.74 | ns |
| TNF-α | Tumor necrosis factor α | ELISA | 1.32 | ns |
| Lymphocytes | Count / µl | FACS | 1.17 | ns |
| NK cells | Count / µl | FACS | 0.80 | * |
| $CD56^{bright}$ NK cells | Count / µl | FACS | 0.78 | ns |
| $CD56^{dim}$ NK cells | Count / µl | FACS | 0.79 | ns |
| T cells | Count / µl | FACS | 0.92 | ns |
| $CD4^+$ T cells | Count / µl | FACS | 0.93 | ns |
| $CD8^+$ T cells | Count / µl | FACS | 0.90 | ns |
| $CD38^+$ $CD4^+$ T cells | Count / µl | FACS | 0.77 | ns |
| $HLA\text{-}DR^+$ $CD4^+$ T cells | Count / µl | FACS | 1.01 | * |
| $HLA\text{-}DR^+$ $CD38^+$ $CD4^+$ T cells | Count / µl | FACS | 1.43 | ns |
| $HLA\text{-}DR^+$ $CD38^+$ $CD8^+$ T cells | Count / µl | FACS | 0.87 | ns |
| $CD38^+$ $CD8^+$ T cells | Count / µl | FACS | 1.10 | ns |
| $HLA\text{-}DR^+$ $CD8^+$ T cells | Count / µl | FACS | 1.26 | ns |
| $CD19^+$ B cells | Count / µl | FACS | 3.42 | ns |
| $CD20^+$ B cells | Count / µl | FACS | 1.65 | ns |

| | | | | |
|---|---|---|---|---|
| Transitional B cells | Count / µl | FACS | 0.89 | ns |
| Naive B cells | Count / µl | FACS | 1.13 | ns |
| Memory B cells | Count / µl | FACS | 3.60 | ns |
| $IgD^+$ memory B cells | Count / µl | FACS | 1.97 | ns |
| $IgD^-$ memory B cells | Count / µl | FACS | 4.37 | ns |
| Plasmablasts | Count / µl | FACS | 0.86 | ns |
| $CD21^{low}$ B cells | Count / µl | FACS | 32.20 | ns |

**Table S5. Variables included in principal component analysis (PCA) in Fig. 6A-B and Fig. S4A-B.** Shown are variables with ratio of means comparing ANA-positive and ANA-negative COVID-19 patients during acute infection (n = 146). P-values were calculated using Mann-Whitney U test. ns, non-significant; *, p < 0.05; **, p < 0.01; ***, p < 0.001; ****, p < 0.0001.

| **Variable name** | **Description** | **Method** | **Mean fold change ANA-pos / ANA-neg** | **p-value** |
| --- | --- | --- | --- | --- |
| Lymphocytes | Count / µl | FACS | 0.93 | ns |
| NK cells | Count / µl | FACS | 0.90 | ns |
| $CD56^{bright}$ NK cells | Count / µl | FACS | 0.87 | ns |
| $CD56^{dim}$ NK cells | Count / µl | FACS | 0.90 | ns |
| T cells | Count / µl | FACS | 0.93 | ns |
| $CD4^+$ T cells | Count / µl | FACS | 0.97 | ns |
| $CD8^+$ T cells | Count / µl | FACS | 0.92 | ns |
| $CD38^+$ $CD4^+$ T cells | Count / µl | FACS | 0.78 | * |
| $HLA\text{-}DR^+$ $CD4^+$ T cells | Count / µl | FACS | 1.11 | ns |
| $HLA\text{-}DR^+$ $CD4^+$ T cells | Count / µl | FACS | 1.10 | ns |
| $CD38^+$ $CD8^+$ T cells | Count / µl | FACS | 0.92 | ns |
| $HLA\text{-}DR^+$ $CD8^+$ T cells | Count / µl | FACS | 1.04 | ns |
| $CD38^+$ $CD8^+$ T cells | Count / µl | FACS | 1.02 | ns |
| $CD19^+$ B cells | Count / µl | FACS | 0.97 | ns |
| $CD20^+$ B cells | Count / µl | FACS | 0.97 | ns |
| Transitional B cells | Count / µl | FACS | 1.28 | ns |
| Naive B cells | Count / µl | FACS | 1.09 | ns |
| $IgD^+$ memory B cells | Count / µl | FACS | 0.74 | *** |
| $IgD^-$ memory B cells | Count / µl | FACS | 0.76 | * |
| Plasmablasts | Count / µl | FACS | 0.71 | ns |
| $CD21^{low}$ B cells | Count / µl | FACS | 0.78 | ns |
| Memory B cells | Count / µl | FACS | 0.75 | * |
| SIgA | S1-specific IgA | ELISA | 1.95 | *** |
| SIgG | S1-specific IgG | ELISA | 1.78 | **** |
| IgG | IgG | ELISA | 1.06 | ns |
| IgG1 | IgG1 | ELISA | 1.12 | * |
| IgG2 | IgG2 | ELISA | 0.96 | ns |
| IgG3 | IgG3 | ELISA | 1.10 | ns |
| IgG4 | IgG4 | ELISA | 1.00 | ns |
| IgM | IgM | ELISA | 0.73 | *** |
| IgA | IgA | ELISA | 1.19 | * |
| IgD | IgD | ELISA | 1.51 | ns |
| IgE | IgE | ELISA | 1.24 | ns |
| IL-1β | Interleukin-β | ELISA | 1.65 | ns |
| IL-2 | Interleukin-2 | ELISA | 0.70 | ns |
| sIL-2Rα | Soluble IL-2 receptor subunit α | ELISA | 1.16 | * |
| IL-5 | Interleukin-5 | ELISA | 1.05 | ns |
| IL-6 | Interleukin-6 | ELISA | 2.28 | * |
| IL-10 | Interluekin-10 | ELISA | 1.44 | *** |
| IL-12 | Interleukin-12 | ELISA | 0.76 | ns |
| IFN-γ | Interferon-γ | ELISA | 0.80 | ns |
| TNF-α | Tumor necrosis factor α | ELISA | 1.32 | *** |
| Age | | History | 1.49 | **** |

**Table S6. Variables included in principal component analysis (PCA) in Fig. 6F-G.** Shown are variables with ratio of means comparing ANA-positive and ANA-negative COVID-19 patients six months after recovery (n = 107). P-values were calculated using Mann-Whitney U test. ns, non-significant; *, $p < 0.05$; **, $p < 0.01$; ***, $p < 0.001$; ****, $p < 0.0001$.

| Marker | Fluorophore | Manufacturer | Clone |
|---|---|---|---|
| IgD | FITC | Beckman Coulter | IA6-2 |
| CD21 | PE | Beckman Coulter | BL13 |
| CD19 | ECD | Beckman Coulter | J3-119 |
| CD27 | PE-Cy7 | Beckman Coulter | 1A4CD27 |
| CD24 | APC | Beckman Coulter | ALB9 |
| CD38 | APC-Alexa Fluor 750 | Beckman Coulter | LS198-4-3 |
| IgM | Pacific Blue | Beckman Coulter | SA-DA4 |
| CD45 | Krome Orange | Beckman Coulter | J33 |
| CD16 | FITC | Beckman Coulter | 3G8 |
| CD56 | PE | Beckman Coulter | N901 |
| CD14 | PE-Cy7 | Beckman Coulter | RMO52 |
| CD4 | APC | Beckman Coulter | 13B8.2 |
| CD8 | Alexa Fluor 700 | Beckman Coulter | B9.11 |
| CD3 | APC-Alexa Fluor 750 | Beckman Coulter | UCHT-1 |
| CD45 | PE-Cy7 | Beckman Coulter | J33 |
| CD3 | ECD | Beckman Coulter | UCHT-1 |
| CD4 | FITC | Beckman Coulter | 13B8.2 |
| HLA-DR | PE | Beckman Coulter | Immu-357 |
| CD38 | PE-Cy5.5 | Beckman Coulter | LS198-4-3 |

**Table S7: Antibody panel for quantification of major lymphocyte subsets by flow cytometry.**
